# *Helicobacter pylori* promotes colorectal carcinogenesis by deregulating intestinal immunity and inducing a mucus-degrading microbiota signature

**DOI:** 10.1101/2022.06.16.22276474

**Authors:** Anna Ralser, Alisa Dietl, Sebastian Jarosch, Veronika Engelsberger, Andreas Wanisch, Klaus Peter Janssen, Michael Vieth, Michael Quante, Dirk Haller, Dirk H. Busch, Li Deng, Raquel Mejías-Luque, Markus Gerhard

## Abstract

**OBJECTIVE:** *H. pylori* infection is the most prevalent bacterial infection worldwide. Besides being the most important risk factor for gastric cancer development, epidemiological data show that infected individuals harbor a nearly two-fold increased risk to develop colorectal cancer (CRC). However, a direct causal and functional connection between *H. pylori* infection and colon cancer is lacking.

**DESIGN:** We infected two *Apc*-mutant mouse models and C57BL/6 mice with *H. pylori* and conducted a comprehensive analysis of *H. pylori*-induced changes in intestinal immune responses and epithelial signatures via flow cytometry, chip cytometry, immunohistochemistry and single cell RNA sequencing. Microbial signatures were characterized and evaluated in germ-free mice and via stool transfer experiments.

**RESULTS:** *H. pylori* infection accelerated tumor development in *Apc*-mutant mice. We identified a unique *H. pylori*-driven immune alteration signature characterized by a reduction in regulatory T-cells and proinflammatory T-cells. Furthermore, in the intestinal and colonic epithelium, *H. pylori* induced pro-carcinogenic STAT3 signaling and a loss of goblet cells, changes that have been shown to contribute - in combination with pro-inflammatory and mucus degrading microbial signatures - to tumor development. Similar immune and epithelial alterations were found in human colon biopsies from *H. pylori*-infected patients. Housing of *Apc*-mutant mice under germ-free conditions ameliorated, and early antibiotic eradication of *H. pylori* infection normalized the tumor incidence to the level of uninfected controls.

**CONCLUSIONS:** Our studies provide evidence that *H. pylori* infection is a strong causal promoter of colorectal carcinogenesis. Therefore, implementation of *H. pylori* status into preventive measures of CRC should be considered.

## INTRODUCTION

*Helicobacter pylori* infection affects more than half of the world’s population and it is a main risk factor for gastric cancer. *H. pylori* induces a number of alterations in the gastric mucosa that together result in neoplastic transformation of the epithelium. Thus, *H. pylori* infection first triggers a complex plethora of immune cascades, directed towards *H. pylori* and orchestrated by the bacterium itself, which originate from priming at the Peyer’s Patches and the mesenteric lymph nodes of the small intestine (1, 2). The major pro-inflammatory response towards *H. pylori* consists of a mixed Th1 and Th17 response (1), and is to a large extent related to the presence and activity of a type 4 secretion system (T4SS) (3), which mediates translocation of the oncogenic and highly immunogenic protein CagA into gastric epithelial cells (4). This leads to chronic inflammation and results in the activation of pro-inflammatory signaling pathways such as activating Nuclear Factor-κB (NF-κB) and signal transducer and activator of transcription 3 (STAT3) signaling, which are major drivers of *H. pylori* induced gastric carcinogenesis (5). However, *H. pylori* has evolved counter mechanisms in order to establish and maintain chronic infection, for example by reprogramming dendritic cells (DCs) to induce regulatory T-cells (Treg) (6, 7), which not only counterbalance the local pro-inflammatory response in the stomach (8), but are also involved in protection from allergic asthma (9). Interestingly, this tolerogenic reprogramming of DCs is partially mediated by CagA, via activation of STAT3 (7). Finally, alterations in gastric microbiota are observed upon infection, which seem to contribute to the deleterious events leading to gastric cancer following *H. pylori* infection (10). This idea is supported by studies using animal models as the insulin-gastrin (INS-GAS) mice, which showed more severe gastric pathology and early development of neoplasia when colonized with *H. pylori* and carrying normal commensal microbiota compared to germ free INS-GAS mice infected with the bacterium (11).

Although *H. pylori* infection is limited to the stomach, accumulating epidemiological data indicate an association between *H. pylori* infection and different extra-gastric diseases (12). Among those, a higher risk of colorectal cancer has been reported to be associated with *H. pylori* infection status (13). However, the mechanisms that could explain this increased risk have not been elucidated.

In our study, we identify *H. pylori-*specific alterations in gut homeostasis that contribute to colorectal carcinogenesis in mouse models of CRC as well as in human samples, and are reversible upon *H. pylori* eradication. These findings provide a basis for assessing *H. pylori* status not only for gastric, but also for colon cancer prevention programmes.

## RESULTS

### *H. PYLORI* PROMOTES INTESTINAL CARCINOGENESIS IN *APC* MOUSE MODELS

To determine whether *H. pylori* infection promotes the development of tumors in the lower gastrointestinal tract, we infected *Apc*^+/min^ and *Apc*^+/1638N^ mice for different time periods (Fig. S1A and Fig. S1B). Unexpectedly, *Apc*^+/min^ mice were highly susceptible to the infection, with only 60% of the mice surviving after 12 weeks of *H. pylori* infection (Fig. 1A). An increased tumor burden in the small intestine and colon was observed in infected *Apc*^+/min^ mice compared to uninfected controls (Fig. 1B and 1C). Similar results were observed in *Apc*^+/1638N^ mice, which not only developed twice as many tumors after infection, but also showed larger tumors in the small intestine (Fig. S1C). Notably, in *Apc*^+/1638N^ mice, colonic tumors were exclusively detected in *H. pylori* infected mice (Fig. S1C). These observations demonstrate that *H. pylori* infection promotes the development of intestinal and colonic tumors in tumor prone mice, while exclusively infecting the stomachs of these mice (Fig S1B).

**Fig. 1:**
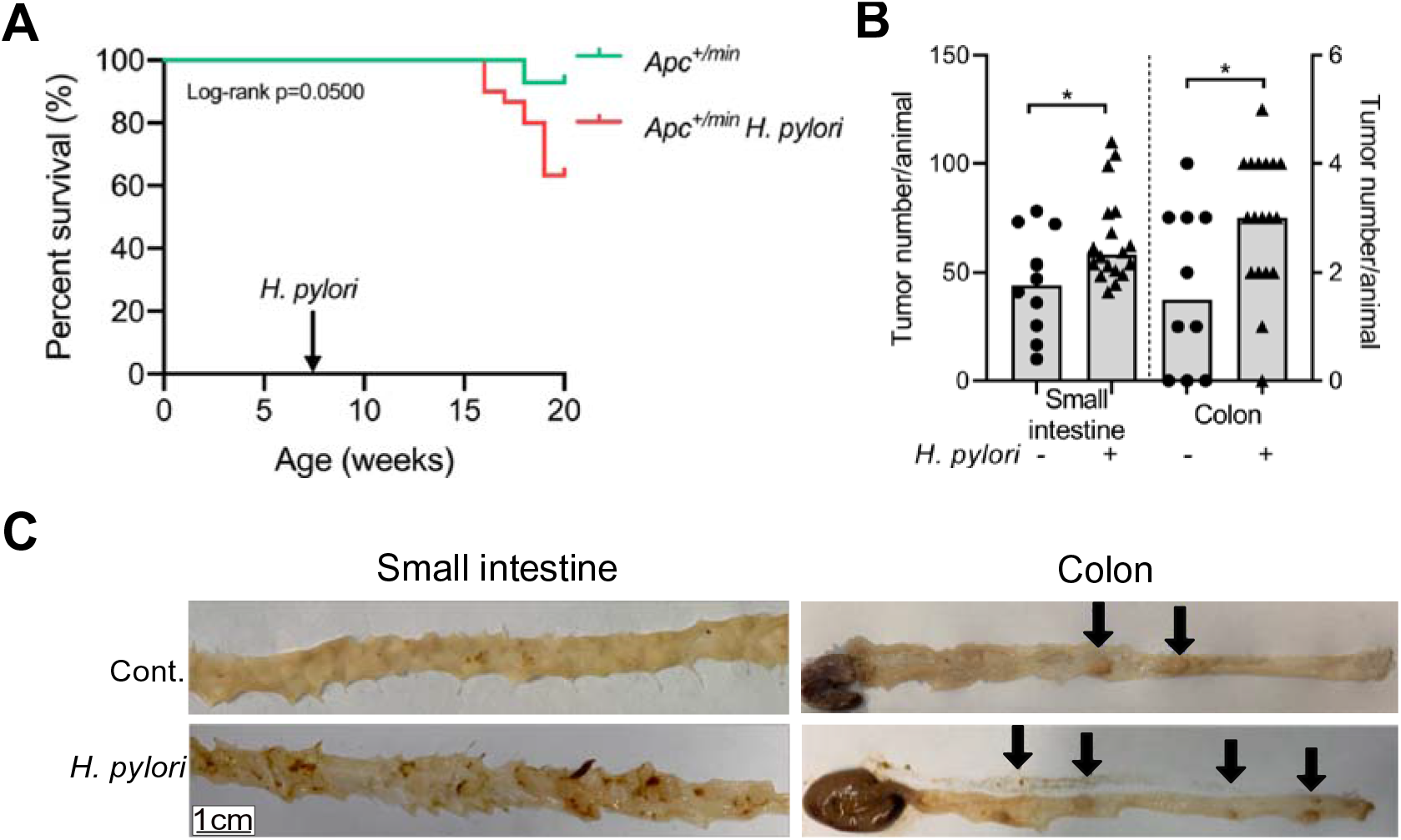
*H. pylori* promotes intestinal carcinogenesis in *Apc* mouse models. (A) Kaplan-Meier survival curve comparing *H. pylori* infected and non-infected *Apc*^+/min^ mice. (B) Tumor counts of *H. pylori* infected (n=18) and non-infected (n=10) *Apc*^+/min^ mice in small intestine and colon. (C) Representative pictures of tumors (arrows) in the small intestine and colon of *H. pylori* infected and non-infected (Cont.) *Apc*^+/min^ mice. Each symbol represents one animal, from 3 independent, pooled experiments. Bars denote median. Statistical significance was determined with Mann-Whitney-U test or unpaired t-test, *p < 0.05.

### *H. PYLORI* INFECTION INDUCES A PRO-INFLAMMATORY RESPONSE IN THE INTESTINE

Manipulation of host’s T cell immune responses characterizes *H. pylori* infection and is one the main mechanisms contributing to gastric carcinogenesis. To assess whether alterations in intestinal immunity could be related to the increased tumor burden observed in infected *Apc* mutant mice, we first analyzed lymphocyte infiltration in the small intestine of *Apc*^+/min^ and *Apc*^+/1638N^ mice upon infection. Recruitment of intraepithelial CD3^+^ T cells to small intestine and colon was increased upon *H. pylori* infection (Fig. 2A and Fig. S2A), wich was also confirmed by flow cytometric analysis of T-cells (Fig. S2B and 2C). Furthermore, this revealed a shift towards more CD8^+^ and less CD4^+^ T cells upon infection (Fig. S2D). In addition, the abundance and protein level of Foxp3^+^ regulatory T (Treg) cells was reduced in the small intestine from infected mice compared to uninfected controls, as detected by flow cytometry (Fig. 2B and Fig. S2E).

**Fig. 2:**
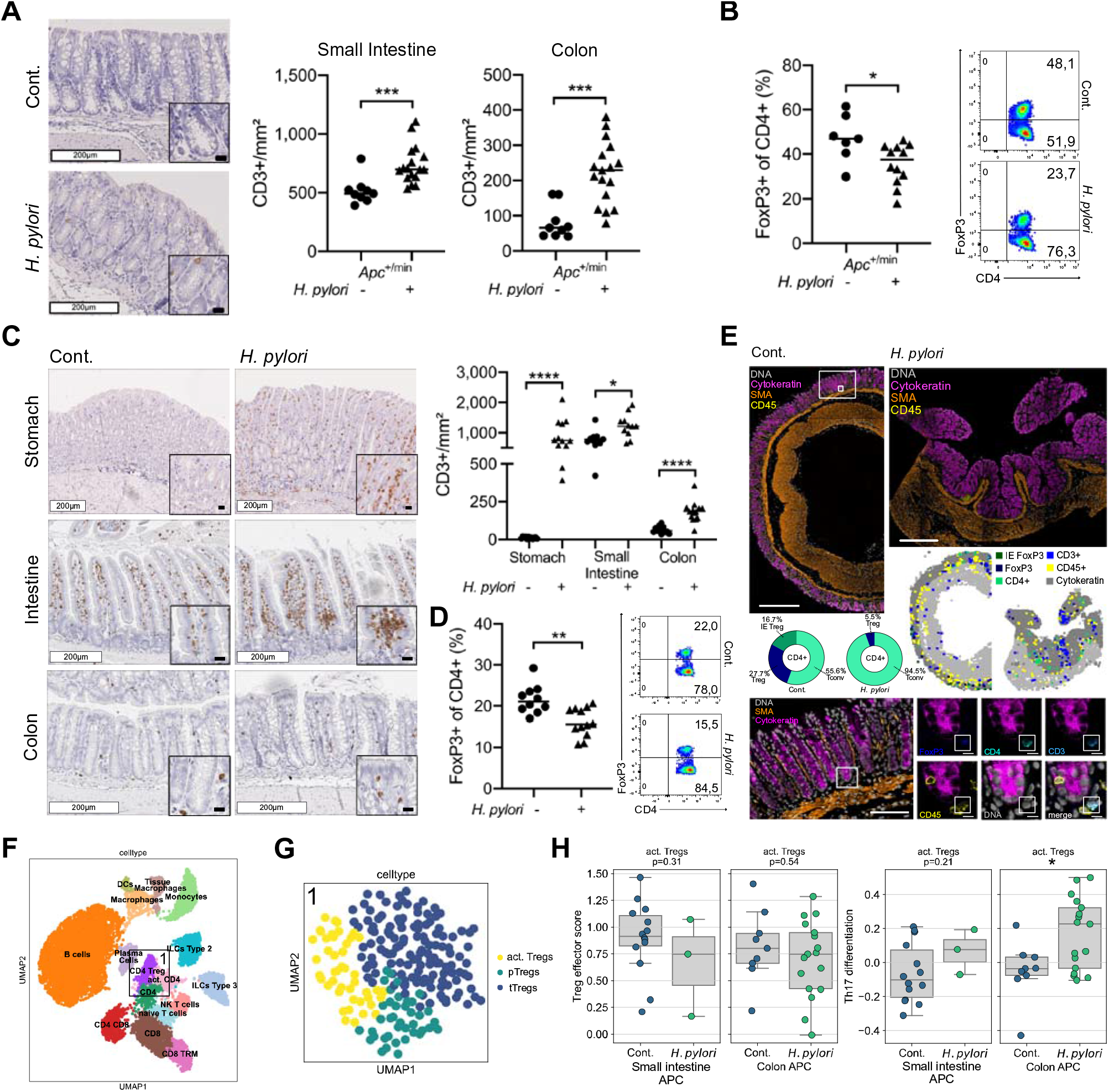
*H. pylori* infection induces a pro-inflammatory response in the intestine. (A) Representative pictures of colonic CD3+ stainings of *H. pylori* infected and non-infected *Apc*^+/min^ mice after 12 weeks of infection are shown. White scale bars correspond to 200 μm, black scale bars to 20 μm. Quantification of positive cells per mm^2^ small intestine and colon tissue is shown. Pooled data of 3 independent experiments. (B) Flow cytometric analysis of intestinal lamina propria lymphocytes isolated from *H. pylori* infected and non-infected *Apc*^+/min^ mice after 12 weeks of infection. Frequency of FoxP3+ positive cells of CD4+ T-cells are shown, gated on live, single cells, CD45+ and CD3+. Pooled data of 2 independent experiments. (C) Representative CD3+ staining of stomach, small intestine and colon tissue sections of *H. pylori* infected and non-infected C57BL/6 mice after 24 weeks of infection. Quantification of intraepithelial cells per mm^2^ is shown. White scale bars correspond to 200 μm, black scale bars to 20 μm. Pooled ata of two independent experiments. (D) Frequency of FoxP3 positive cells of CD4+ T-cells are shown. Cells were gated on live, single cells, CD45+ and CD3+. Pooled data of 2 independent experiments. (E) Overview of colon tissue stained with multiplexed chip cytometry. Automatic image processing of multiplexed chip cytometry on colon tissue determines CD4+ T-cell properties in *H. pylori* positive and negative C57BL/6 mice. Frequencies of conventional T-cells (Tconv), regulatory T-cells (Treg) and intraepithelial regulatory T-cells (IE Treg) are shown. Scale bar in overview corresponds to 500μm. Representative picture of colon tissue stained with multiplexed chip cytometry. FoxP3+ cell, defined by intranuclear FoxP3+, CD4+ CD3+ and CD45+ staining. Large scale bar corresponds to 100μm, small scale bars to 10μm. Representative data of one experiment. (F) Annotated immune cells plotted as UMAP, clusters for further analysis are highlighted (1=CD4 Treg, 2=CD8 and CD8 TRM). (G) Unsupervised clustering and annotation of Treg cluster as UMAP, n=217 cells. activated Tregs = act. Tregs, pTregs =peripherally induced Tregs, tTregs = thymically derived Tregs. (H) Gene set score of Treg effector genes and Th17 differentiation genes, comparing activated Treg cells from small intestine and colon of *H. pylori* infected and non-infected *Apc*^+/min^ (APC) mice. Statistical significance was determined with Kruskal-Wallis test. Each symbol represents one animal/single cell, pooled from at least 2 independent experiments (n=6-10 mice/group/experiment) or 2 mice/group for single-cell data. Bars denote median. Unless otherwise specified, statistical significance was determined with student’s t-test in case of normal distribution, otherwise by Mann-Whitney U test, *p < 0.05, **p < 0.01, ***p < 0.001, ****p < 0.0001.

To further explore the underlying mechanisms promoting intestinal tumorigenesis upon *H. pylori* infection independently from tumor-prone backgrounds, we infected wild type C57BL/6 mice (wt) and analyzed immune responses (Fig. S2F). An increased number of intraepithelial CD3^+^ T cells was also observed in the small intestine as well as in the colon of *H. pylori* infected wt mice compared to uninfected controls (Fig. 2C). Contrasting the balanced immune phenotype observed in the stomach (Fig. 2C and Fig. S2G), this was accompanied by a reduction in Foxp3^+^ Treg cells (Fig. 2D and Fig. S2G). Multiplexed ChipCytometry corroborated an overall reduction of Treg cells, and additionally revealed their compartmentalization within the lamina propria in infected colonic tissue (Fig. 2E).

We next confirmed the specificity of these T cells for *H. pylori* by restimulating lamina propria CD4^+^T cells with *H. pylori* lysate and measuring the release of the pro-inflammatory cytokine IL-17A, which has been previously described to be one of the main players in the immune response to *H. pylori* (1). A specific IL-17A/CD4^+^T cell response was observed in infected C57BL/6 and *Apc*^+/min^ mice (Fig. S2H).

To characterize in depth the specific intestinal immune response elicited by gastric *H. pylori*, we investigated the immune cell compartment on a single cell level by performing 10X single-cell RNA sequencing (scRNAseq). We isolated and sorted single CD45^+^ immune cells of intestinal and colonic tissue from *Apc*^+/min^ mice and littermate wild type controls that had been infected for 12 weeks with *H. pylori*, and compared them to non-infected controls (Fig. S2I). Unsupervised clustering identified 16 clusters according to their transcriptional profiles, which are visualized using Uniform Manifold Approximation and Projection (UMAP) (14) (Fig 2F and Fig. S2J) and were annotated based on known marker genes (Fig. S2K).

To further characterize the Treg cell compartment, we subclustered and annotated Treg cells, which resulted in three subclusters: activated Treg cells (act. Tregs); peripherally derived Treg cells (pTregs), characterized by high RORyt expression; and thymus derived Treg cells (tTregs), characterized by GATA3 expression (15, 16) (Fig. 2G and Fig. S2L). We then computed a Treg effector score (17) (Fig. 2H), and found significantly increased Th17 differentiation genes in infected act. Tregs (Fig. 2H), indicating that *H. pylori* infection reprograms Treg cells into potentially pathogenic Foxp3^+^ IL-17A^+^ T cells.

Finally, to understand cell dynamics of T cells in infected mice, we calculated RNA velocity vectors, which predict future states of individual cells based on ratios of spliced and unspliced mRNAs (18). In line with our previous findings, when looking at the CD4 clusters, it was apparent that less CD4 cells were projected towards CD4 Treg cells in infected *Apc* mice (Fig. S2M).

In summary, our results show that *H. pylori* infection induces a *H. pylori* specific pro-inflammatory immune response in the small intestine and colon of infected mice, that is characterized by loss of regulatory T cells and their differentiation into Foxp3^+^ IL-17A^+^T cells.

### ACTIVATION OF CARCINOGENIC SIGNALING PATHWAYS AND LOSS OF GOBLET CELLS CHARACTERIZE THE INTESTINAL EPITHELIAL RESPONSE TO *H. PYLORI* INFECTION

Considering the alterations induced by *H. pylori* in intestinal immune cells independently of *APC* mutations in wild type mice, we analyzed the effect on signaling pathways putatively mediating the pro-carcinogenic effects of *H. pylori* infection in the epithelium. Therefore, we assessed transcriptomic profiles of EPCAM^+^ epithelial cells from *Apc*^+/min^ mice in our scRNAseq data (Fig. S2I). Unsupervised clustering revealed 15 clusters according to their transcriptional profiles, which were visualized as UMAP (Fig. 3A and Fig. S3A) and annotated based on known marker genes (Fig. S3B).

**Fig. 3:**
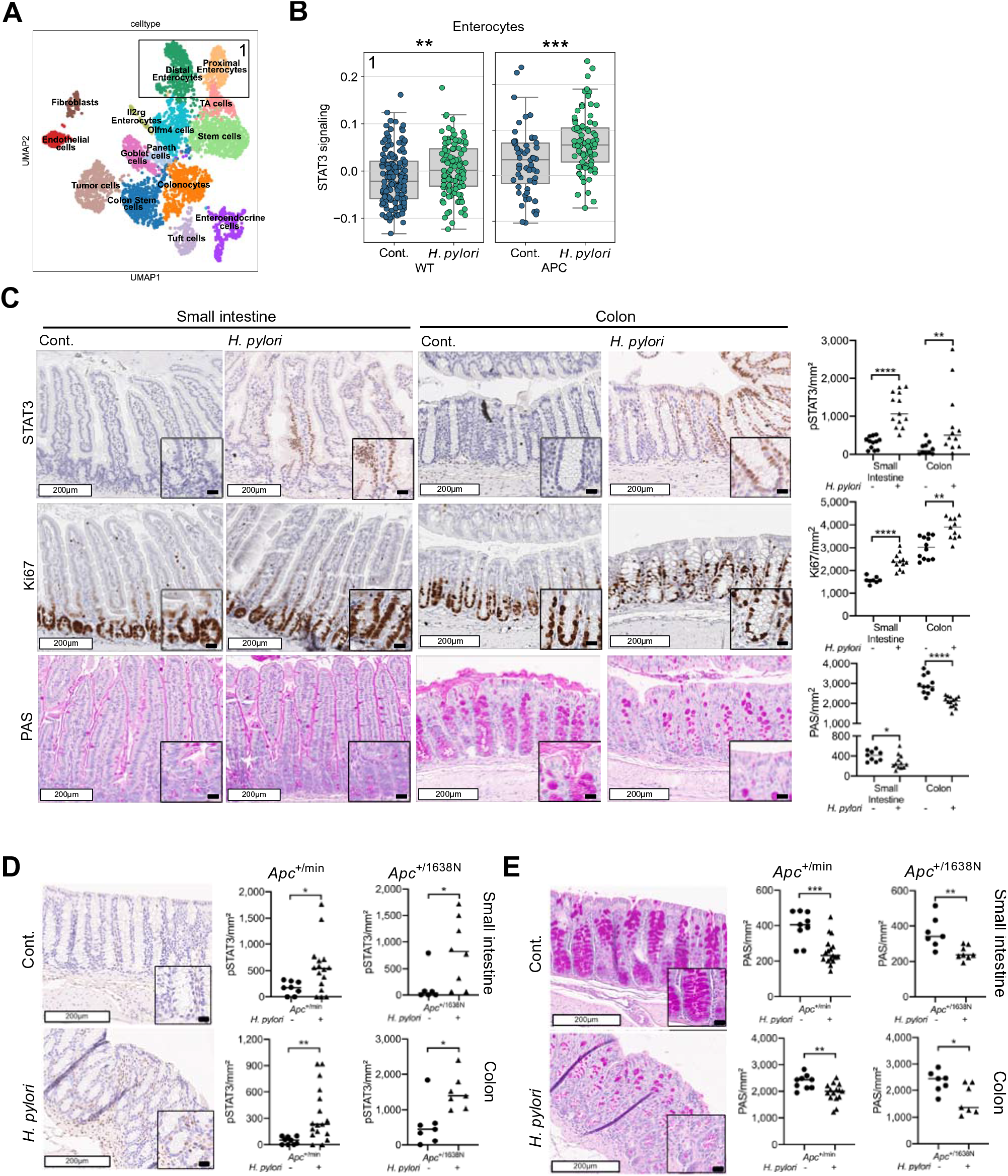
Activation of carcinogenic signaling pathways and loss of goblet cells characterize the intestinal epithelial response to *H. pylori* infection. (A) Annotated epithelial cells after unsupervised clustering plotted in UMAP space, n=2 mice per group, n= 4249 cells. Clusters for further analysis are highlighted (1= Enterocytes) (B) Gene set score of STAT3 signaling genes, comparing intestinal enterocytes from *H. pylori* infected and non-infected *Apc*^+/+^ (WT) and *Apc*^+/min^ (APC) mice. Statistical significance was determined with Kruskal-Wallis test. (C) Representative pSTAT3, Ki67 and PAS staining of small intestine and colon tissue of *H. pylori*-infected and non-infected C57BL/6 mice. Quantification of positive cells per mm^2^ is shown. White scale bars correspond to 200 μm, black scale bars to 20 μm. Pooled data of 2 independent experiments. (D) Representative pictures of colonic pSTAT3 staining of *H. pylori* infected and non-infected *Apc*^+/min^ and *Apc*^+/1638N^ mice are shown. White scale bars correspond to 200 μm, black scale bars to 20 μm. Quantification of positive cells per mm^2^ are shown. Pooled data of 3 independent experiments. (E) Representative pictures of colonic PAS staining of *H. pylori* infected and non-infected *Apc*^+/min^ and *Apc*^+/1638N^ mice are shown. White scale bars correspond to 200 μm, black scale bars to 20 μm. Quantification of positive cells per mm^2^ are shown. Pooled data of 3 independent experiments. Each symbol represents one animal/single cell, pooled from at least 2 independent experiments (n=6-10 mice/group/experiment) or 2 mice/group for single-cell data. Bars denote median. Unless otherwise specified, statistical significance was determined with student’s t-test in case of normal distribution, otherwise by Mann-Whitney U test, *p < 0.05, **p < 0.01, ***p < 0.001, ****p < 0.0001.

Pseudo-spatial distribution of epithelial cells along the crypt-villus axis were computed to confirm correct annotation of cell types (19, 20) (Fig. S3C).

Here, we specifically explored signaling pathways associated with CRC initiation and development, namely STAT3 and NF-κB. Notably, these pathways also orchestrate key inflammatory mechanisms in inflammation-driven colon cancer and have been extensively related to *H. pylori* infection (21–23). We computed a score of genes involved in the Jak-STAT signaling pathway, which revealed significantly higher scores in enterocytes and stem cells of wild type and enterocytes of *Apc*^+/min^ mice upon *H. pylori* infection (Fig. 3B, Fig. S3D). Similarly, when assessing NF-κB signaling in enterocytes, higher scores were observed upon *H. pylori* infection (Fig. S3E).

As it has been shown that activation of epithelial STAT3 favors recruitment of lymphocytes, while inhibiting infiltration of Treg cells in the colon (24), we confirmed hyperactivation of STAT3 in tissue samples from *H. pylori* infected wild type (Fig. 3C) and *Apc* mutant mice (Fig. 3D), which was accompanied by enhanced proliferation as detected by Ki67 staining (Fig. 3C).

Given that a functional intestinal barrier is depending on mucus replenishment by goblet cells, we next assessed their status by Periodic acid Schiff (PAS) staining. We observed reduced number of mucus producing cells in the small intestine and in the colon of *H. pylori* infected wild type (Fig. 3C) and *Apc* mutant mice (Fig. 3E) compared to uninfected controls. To explore in depth the effects of *H. pylori* infection on the goblet cells, we clustered and annotated goblet cells in our scRNAseq data set based on goblet cell differentiation markers (25). This revealed immature, characterized by high expression of Tff3; intermediate, highly expressing Oasis; and terminal goblet cells, with high expression of Muc2 and Klf4 (Fig. S3F and Fig. S3G). Maturational states were distinctly affected by *H. pylori* infection, with a switch to less differentiated goblet cells (Fig. S3H). To assess goblet cell functionality, we assessed the expression of antimicrobial peptide genes *Reg3b* and *Reg3g*, which are known to play a role in response to pathogens and inflammation (26). We found them to be reduced upon *H. pylori* infection (Fig. S3I). These findings are consistent with a compromised intestinal barrier integrity induced by *H. pylori* infection, independent of *APC* status.

To explain the absolute loss of goblet cells we observed in infected mice, we studied cellular dynamics of goblet cells by means of RNA velocities. In the colon, we observed less directionality from the stem cell cluster towards the goblet cell cluster, and at the same time a higher projection towards the colonocyte cluster upon *H. pylori* infection (Fig. S3J), in contrast to the small intestinal goblet cell cluster, where cell dynamics seem to be restricted to the goblet cell cluster itself. When assessing the expression of Atoh1, which is known to drive terminal differentiation into the secretory lineage (27), in stem cells of both small intestine and colon, a signficiantly lower expression was found upon *H. pylori* infection (Fig. S3K). These findings indicate a skewed differentiation of stem cells rather into unspecialized colonozytes than into goblet cells.

Together, these results indicate that *H. pylori* induces carcinogenic signaling pathways and has a detrimental impact on mucus producing goblet cells in the small intestine and colon of wt and *Apc* mutant mice.

### *H. PYLORI* INFECTION FAVORS THE PRESENCE OF MUCUS-DEGRADING MICROBIOTA

Microbiota alterations and aberrant presence of certain bacterial species in the small intestine have been related to the development of CRC (28). This could be an additional mechanism by which *H. pylori* contributes to intestinal carcinogenesis, since *H. pylori* infection has been shown to alter microbiota signatures (29). Therefore, we assessed to which extent *H. pylori* infection influenced small intestinal and colonic microbial composition by performing 16S RNA sequencing. While we found significantly increased abundance of *Helicobacter* spp. in the stomach, we did not detect *H. pylori* in intestine and colon (Fig. S4A). When comparing the microbiota in caecum and colon of infected and non-infected mice via taxonomic profiling, we observed apparent changes at phylum level upon *H. pylori* infection (Fig. 4A). Furthermore, we found signs of decreased α-diversity in small intestine upon *H. pylori* infection (Fig. S4B) as well as significantly different β-diversity in caecum, stool and small intestine between non-infected and infected mice (Fig. S4C). Differential abundance testing revealed *Akkermansia* spp. to be enriched in infected wt mice (Fig. S4D, Fig. 4B). When exploring the data for further species sharing the mucus-degrading characteristics of *Akkermansia* spp., we found an increase in *Ruminococcus* spp. (Fig. 4B and Fig. S4D). The abundance of both species was also higher in *Apc* mutant mice upon *H. pylori* infection (Fig. 4C and 4D).

**Fig. 4:**
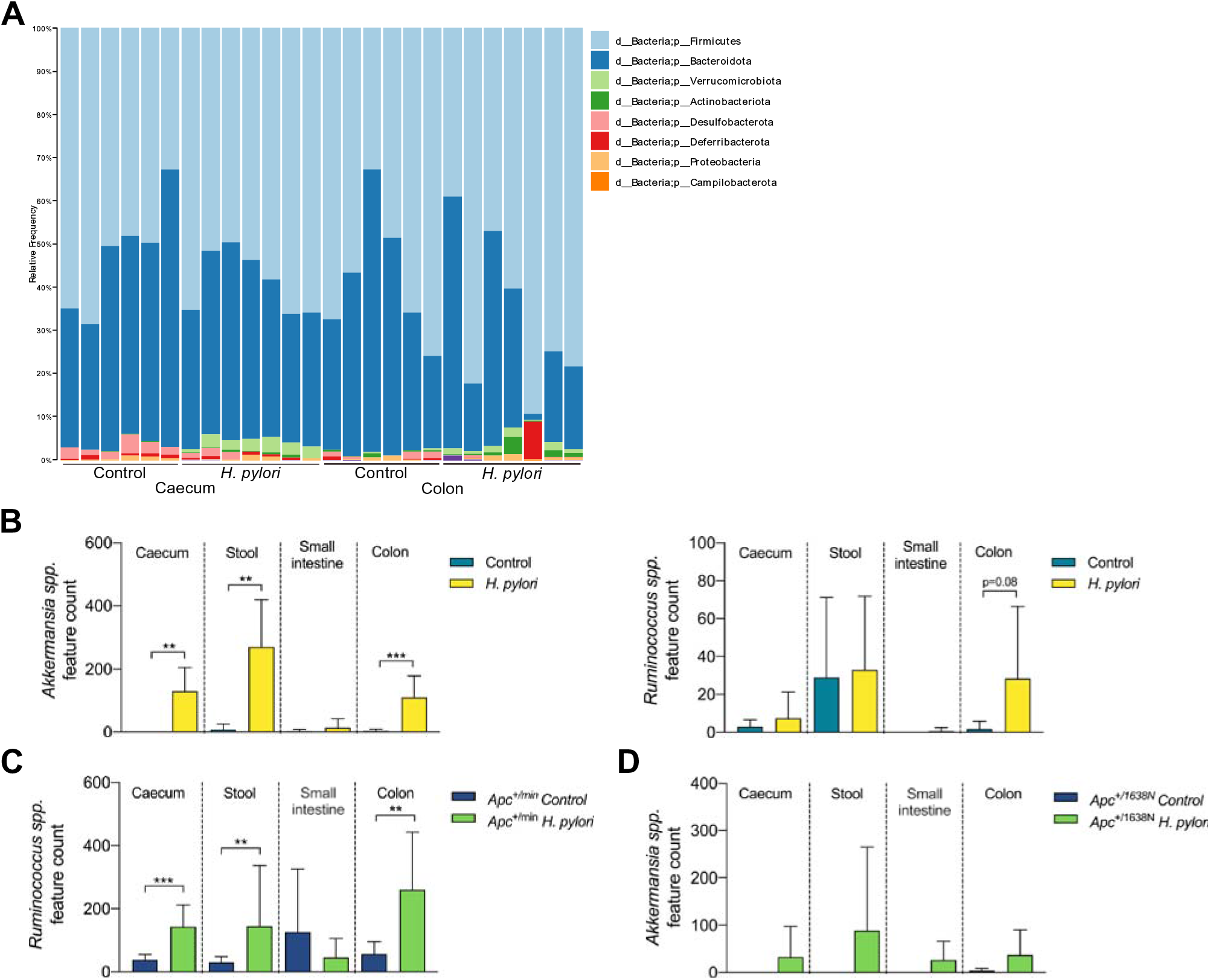
*H. pylori* infection favors the presence of mucus-degrading microbiota. (A) Relative taxonomic frequencies on phyla level in 16S rRNA sequenced caecum and colon samples of *H. pylori* infected and non-infected C57BL/6 mice (n=6-8mice/group). (B) Feature counts (ASVs) of *Akkermansia spp.* and *Ruminococcus spp.* in ceacum, stool, small intestine and colon of *H. pylori* infected and non-infected C57BL/6 mice (n=6-8mice/group). (C) Feature counts (ASVs) of *Ruminococcus spp.* of ceacum, stool, small intestine and colon of *H. pylori* infected and non-infected *Apc*^+/min^ mice (n=4-5mice/group). (D) Feature counts (ASVs) of *Akkermansia spp.* of ceacum, stool, small intestine and colon of *H. pylori* infected and non-infected *Apc*^+/1638N^ mice (n=4-5mice/group). Data of one representative experiment of 2-3 independent experiments, shown as bars with mean and standard deviation (SD). Statistical significance was determined with Mann-Whitney U test, **p < 0.01, ***p < 0.001.

Together, *H. pylori* alters the microbiota of the lower gastrointestinal tract and induces distinct mucus-degrading signatures in both wt and *Apc* mutant mice.

### *H. PYLORI* INDUCED COLORECTAL CARCINOGENESIS IS PREVENTED BY ERADICATION

The interplay between a pro-inflammatory response and activation of pro-carcinogenic signaling, accompanied by alterations in microbiota characterized *H. pylori*-driven intestinal tumorigenesis. To dissect the contribution of inflammation in the absence of microbiota, we infected *Apc*^+/1638N^ mice under germ-free conditions (Fig. S5A and 5B). We observed similar immune alterations as in specific-pathogen free (SPF) mice, namely increased CD3^+^ T cell infiltration and reduction of Treg cells in small intestine and colon (Fig. 5A, Fig. S5D and 5B). In contrast, germ-free mice barely showed activation of STAT3 signaling, and no reduction of goblet cells upon *H. pylori* infection (Fig. 5C and 5D, Fig. S5D). Finally, we still observed a higher tumor number in *H. pylori* infected germ-free mice, albeit not significant (Fig. 5E). In order to ultimately assess the contribution of *H. pylori* induced changes in microbiota to intestinal carcinogenesis, we performed a stool transfer experiment. Stool was obtained from 4 different groups, namely SPF non-infected and *H. pylori* infected *Apc*^+/1638N^ and *Apc*^+/+^ mice, respectively, and transferred into germ-free *Apc*^+/1638N^ mice (Fig. S5C). Higher tumor numbers in stool recipients from *H. pylori* infected mice were found, which was already evident in *Apc*^+/+^ mice and further enhanced in an *Apc*^+/1638N^ background (Fig. 5F). This indicates a strong contribution of *H. pylori-*induced changes in microbiota to the tumor phenotype and suggests, that *H. pylori-*induced carcinogenesis in the small intestine and colon is a multifactorial process involving the interplay of the pro-inflammatory immune response, alterations in microbiota and procarcinogenic signaling. Therefore, we next sought to determine whether eradication of *H. pylori* infection could abrogate the carcinogenic process, by treating the mice with a triple therapy regimen consisting of clarithromycin, metronidazole and omeprazole (30), reflecting the “Italian triple therapy” regimen also used in infected patients to eradicate *H. pylori* (Fig. 5G and Fig. S5E). Importantly, we found that after antibiotic eradication, tumor burden was at the same levels as in uninfected controls (Fig. 5H). To delineate that the underlying changes in the intestinal immune response were directly induced by *H. pylori* infection and independent of mutated *Apc*, we analyzed the effect of *H. pylori* eradication on C57Bl/6 mice (Fig. 5G), and found a lower CD3^+^ T cell infiltration in the stomach (Fig. S5F), small intestine and colon (Fig. 5I, Fig. S5G) compared to infected mice at 4 and 12 weeks post eradication. The percentage of Treg cells was initially reduced in eradicated mice 4 weeks after treatment, while 12 weeks post-treatment, the percentage of FoxP3^+^ T cells was observed to recover (Fig. 5J). A specific IL-17A/CD4^+^T cell response was observed in infected mice, which was initially lost after eradication therapy, but then reappeared after longer recovery time (Fig. 5K), supporting the specificity of the response to *H. pylori*, based on the given antigen encounter and response also in eradicated mice. Importantly, the clearance of infection resulted in normalization of STAT3 activation and the number of PAS positive cells (Fig. 5L and Fig. S5G), confirming that *H. pylori* is specifically responsible for these changes.

**Fig. 5:**
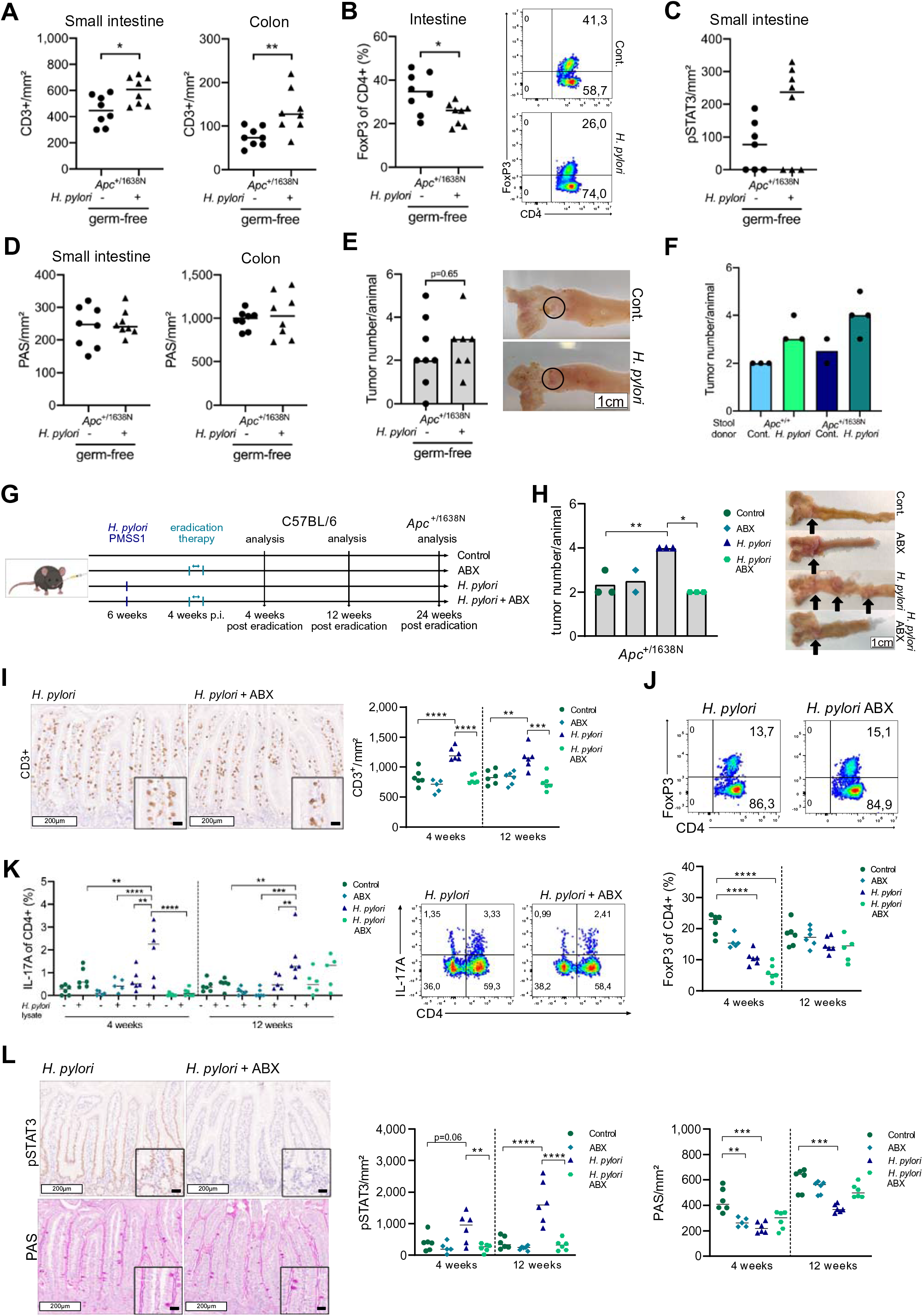
*H. pylori* induced intestinal carcinogenesis is prevented by eradication. (A) Quantification of small intestinal and colonic intraepithelial CD3+ cells per mm^2^ of *H. pylori* infected and non-infected germ-free *Apc*^+/1638N^ mice is shown. Pooled data of two independent experiments (n= 8 mice/group). (B) Frequencies of FoxP3+ cells of CD4+ T-cells, gated on live, single cells, CD45+ and CD3+ of *H. pylori* infected and non-infected germ-free *Apc*^+/1638N^ mice are included. Representative pseudocolor plots are shown. Pooled data of two independent experiments (n= 8 mice/group). (C) Quantification of intestinal intraepithelial pSTAT3+ cells per mm^2^ of *H. pylori* infected and non-infected germ-free *Apc*^+/1638N^ mice is shown. Pooled data of two independent experiments (n= 8 mice/group). (D) Quantification of intestinal and colonic PAS+ cells per mm^2^ of *H. pylori* infected and non-infected germ-free *Apc*^+/1638N^ mice is shown. Pooled data of two independent experiments (n= 8 mice/group). (E) Intestinal tumor counts of *H. pylori* infected and non-infected germ-free *Apc*^+/1638N^ mice and representative pictures of tumors (circled) are shown (n=8 mice/group). (F) Intestinal tumor counts of stool transfer experiments from non-infected and *H. pylori* infected *Apc*^+/+^ or *Apc*^+/1638N^ mice, respectively, (stool donors) into germ-free *Apc*^+/1638N^ mice (stool recipients) are shown. Data of one experiment (n=2-4 mice/group). (G) Experimental setup of *H. pylori* eradication therapy of C57BL/6 and *Apc*^+/1638N^ mice. (H) Tumor counts of non-infected, antibiotically treated, *H. pylori* infected and *H. pylori* eradicated *Apc*^+/1638N^ mice and representative pictures of tumors in the small intestine. Data of one experiment (n= 2-3 mice/group). (I) Representative pictures of CD3+ stainings from small intestinal tissue of *H. pylori* infected and eradicated C57BL/6 mice are shown. White scale bars correspond to 200 μm, black scale bars to 20 μm. Quantification of positive cells per mm^2^ is shown. Data of one experiment (n= 5-6 mice/group). (J) Flow cytometric analysis of intestinal lamina propria lymphocytes reveals frequency of FoxP3+ CD4+ T-cells, gated on live, single cells, CD45+ and CD3+. Data of one experiment (n= 5-6 mice/group). (K) Flow cytometric analysis of intestinal lamina propria lymphocytes reveals IL-17A release of CD4+ T-cells, restimulated with whole *H. pylori* lysate. Cells are gated on live, single cells, CD45+ and CD3+. Data of one experiment (n= 5-6 mice/group). (L) Representative pictures of pSTAT3 and PAS staining from intestinal tissue of H. pylori infected and eradicated C57BL/6 mice are shown. White scale bars correspond to 200 μm, black scale bars to 20 μm. Quantification of positive cells per mm^2^ is shown. Data of one experiment (n= 5-6 mice/group). Each symbol represents one animal. Bars denote median. Statistical significance between two groups was determined with student’s t-test in case of normal distribution and otherwise with Mann-Whitney U test. Among more than 2 groups, ordinary one-way ANOVA with Tukey’s multiple comparisons test was applied in case of normal distribution, otherwise Kruskal-Wallis-Test with Dunn’s multiple comparisons test, *p < 0.05, **p < 0.01, ***p < 0.001, ****p < 0.0001.

In summary, our results demonstrate that *H. pylori* directly enhances colon carcinogenesis by shaping intestinal and colonic immune responses and inducing profound changes in intestinal/colonic microbiota and epithelial homeostasis. Eradication of *H. pylori* infection prevents its tumor-promoting effects also in the colon, providing a possible additional strategy to reduce CRC burden.

### *H. PYLORI* ALTERS COLONIC HOMEOSTASIS IN HUMAN

Our mouse models showed that *H. pylori* affects intestinal and colonic homeostasis at different cellular and molecular levels, which can ultimately enhance tumor development. To determine whether these effects were also observed in humans, we analyzed immune signatures in a cohort of 154 human colon tissue samples. Based on immune responses and histology of the stomach, we stratified samples according to *H. pylori* status into currently (actively) infected and eradicated patients. We found that *H. pylori*-actively infected as well as eradicated individuals showed higher infiltration of CD3^+^ T cells in the colon compared to uninfected subjects (Fig. 6A). Using endoscopy-derived colon biopsies, we further characterized T cell responses by flow cytometry, which revealed tendencies towards more CD3^+^ T cells in the colonic mucosa of currently infected patients (Fig. S6A). In contrast, CD4^+^ and CD8^+^ subsets were not affected by *H. pylori* status (Fig. S6B). Notably, the number of FoxP3^+^ cells in the colonic mucosa was lowest in the currently infected group, whereas eradicated patients seem to level with negative controls (Fig. 6C). The overall loss of Tregs was confirmed via ChipCytometry (Fig. 6B), which additionally showed that intraepithelial localization of Tregs is almost lost in colon samples from *H. pylori* infected individuals (Fig. 6B and Fig. S6C).

**Fig. 6:**
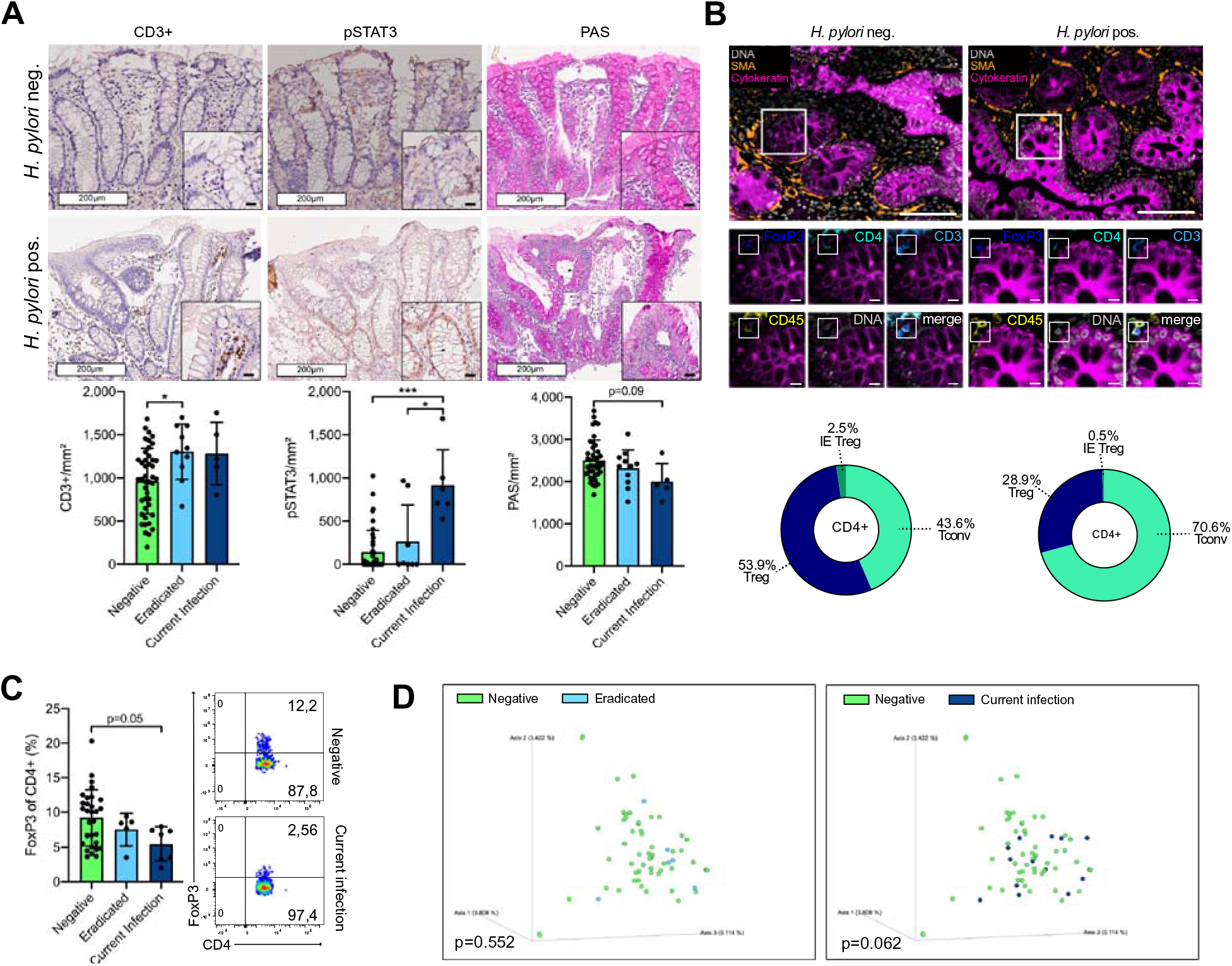
*H. pylori* alters colonic homeostasis in human. (A) Representative HE, CD3+, pSTAT3 and PAS pictures of colonic tissue from *H. pylori* currently infected, eradicated and non-infected patients are shown. White scale bars correspond to 200 μm, black scale bars to 20 μm. Quantification of total CD3+, intraepithelial STAT3 and PAS positive cells per mm^2^ are shown. (B) Representative pictures of human colon tissue stained by multiplexed chip cytometry. FoxP3+ cell, defined by intranuclear FoxP3+, CD4+, CD3+ and CD45+ staining are shown for *H. pylori* negative and positive tissue. Large scale bar corresponds to 100μm, small scale bar to 10μm. Automatic image processing of multiplexed chip cytometry on colon tissue determines CD4+ T-cell properties in *H. pylori* positive and negative individuals: frequencies of conventional T-cells (Tconv), regulatory T-cells (Treg) and intraepithelial regulatory T-cells (IE Treg) are shown. (C) Flow cytometric analysis of colon biopsies from *H. pylori* currently infected, eradicated and non-infected patients were conducted. Frequencies of FoxP3+ cells of CD4+ T-cells, gated on live, single cells, CD45+ and CD3+ are shown and representative pseudocolor plots of *H. pylori* currently infected and negative individuals are included. (D) Bray-Curtis dissimilarity depicting beta-diversity between *H. pylori* infected and eradicated as well as *H. pylori* infected and non-infected patients. Statistical significance was determined with PERMANOVA. Each symbol represents one patient, shown as bars with mean and standard deviation (SD). Statistical significance was determined with ordinary one-way ANOVA with Tukey’s multiple comparisons test in case of normal distribution, otherwise by Kruskal-Wallis-Test with Dunn’s multiple comparisons test. *p < 0.05, **p < 0.01, ***p < 0.001.

We next focused on the epithelial compartment and, in concordance with our findings in mice and our eradication experiments, found a higher number of p-STAT3 positive epithelial cells and a concomitant loss of mucus producing cells in the colon of currently infected subjects, which was attenuated in eradicated patients (Fig. 6A).

Finally, we assessed microbial changes in stool of patients and found a difference in β-diversity between actively *H. pylori* infected and negative patients (p=0.062), but not between *H. pylori* eradicated and negative patients (p=0.552) (Fig. 6D). In contrast, we neither detected significant changes in alpha-diversity (Fig. S6D) nor in Firmicutes to Bacteroidetes ratio (Fig. S6E) between the 3 groups. Comparative microbiome profiling revealed *Prevotellaceae* and *Peptostreptococcales*, which have been associated with CRC, to be differentially abundant in *H. pylori* positive patients (Fig. S6F and Fig. S6G).

These results confirm that the immune and epithelial signatures identified in mouse models upon *H. pylori* infection are also observed in humans, and are accompanied by changes in microbiota compositions, which can further contribute to colon carcinogenesis. Furthermore, the attenuated phenotype observed in *H. pylori* eradicated patients further supports *H. pylori* status as an independent risk factor for CRC and simultaneously offers an option for CRC prevention for those patients at risk.

## DISCUSSION

Although selectively colonizing the stomach, chronic *H. pylori* infection is associated with several extragastric diseases (31). Epidemiological data indicates an association between *H. pylori* infection and a higher risk and aggressiveness of colorectal cancer (CRC), with an odds ratio (OR) of 1.9 (32), an OR higher than for most other known risk factors, such as smoking, alcohol and BMI (32). However, these epidemiological data have not yet been confirmed experimentally, and a molecular mechanism by which *H. pylori* may promote CRC remained elusive. We employed *Apc* mutant mouse lines (*Apc*^+/min^ and *Apc*^+/1638N^) as surrogate models for human CRC, and observed a nearly two-fold increase in tumor numbers in mice infected with *H. pylori*, which coincides with the OR observed in epidemiological studies. Remarkably, this increase was not only observed in the small intestine, where both models usually show most tumors, but was especially evident in the colon. This prompted us to decipher the potential mechanisms driving *H. pylori*-induced carcinogenesis in the small intestine and colon.

The effects of *H. pylori* infection on other organs are best understood for the lung, where chronic *H. pylori* infection imposes a regulatory immune signature that protects from asthma disease (33). In contrast to these observations, we observed an *H. pylori* antigen-specific pro-inflammatory Th17 mediated response in the small intestine and colon, which was not balanced by an increase in Treg cells, as it occurs in the stomach or lung. The immune response mounted towards *H. pylori* originates from Peyer’s patches in the gut (2), which may explain why *H. pylori* specific T cells also homed to intestinal and colonic mucosal sites. Interestingly, IL-17 was not only found to be increased in *H. pylori* positive patients with gastritis and gastric cancer (34), but also in CRC, where Th17 signatures, including RORC, IL17, IL23 and STAT3, were linked to poorer prognosis (35). Still, it was surprising to observe a loss of intestinal Treg cells, which also contrasts the balanced immune response usually observed in the stomach upon *H. pylori* infection. Additionally, we found that in the lower GI tract, Tregs were reprogrammed to upregulate Th17 differentiation markers. Murine and human studies have demonstrated that Treg cells can be reprogrammed to a distinct population, Foxp3^+^/IL17^+^T cells, phenotypically and functionally resembling Th17 cells (36). Particularly in CRC, the presence of Foxp3^+^/IL17^+^T cells has been reported to be increased in the mucosa and peripheral blood of chronic colitis patients as well as in colorectal tumors (37). Foxp3^+^/IL17^+^ cells were shown to promote the development of tumor-initiating cells by increasing the expression of several CRC-associated markers such as CD44 and epithelial cell adhesion molecule EPCAM in bone marrow-derived mononuclear cells (38). Thus, the pro-inflammatory Th17 response elicited by *H. pylori*, especially the differentiation of Treg cells to a Th17 phenotype, may constitute one of the major mechanisms enhancing tumor development. This is in line with literature showing that altered T cell homeostasis is a key event during colorectal carcinogenesis, not only driving tumor development and progression, but also determining treatment response of CRC patients (39).

Mutations in the gene encoding adenomatous polyposis coli (*APC*) are the most frequent driver mutations leading to sporadic CRC, together with mutations in *TP53* and *KRAS* (40, 41). Pro-inflammatory and proliferative signaling pathways such as STAT3, NF-κB and WNT signaling, activated by signals derived from epithelial and immune cells, drive chronic inflammation, a known mechanism contributing to CRC (23, 42). CRC risk is markedly increased in patients with chronic inflammatory bowel disease (IBD), with the risk rising with the duration of disease, from 8.3% after 20 years, to 18.4% after 30 years (43). Mechanistically, besides immune signaling by Th17 cells, activation of pro-inflammatory signaling pathways as well as altered microbiota, contribute to the pathogenesis of colitis-associated cancer (44).

The strong pro-inflammatory response induced locally by *H. pylori* in the small intestine was accompanied by the activation of NF-κB and STAT3 pathways. Activation of STAT3 signaling has been strongly related to tumor initiation, development and progression, while levels of activated STAT3 in the tissue correlate with tumor invasion, TNM stage and reduced overall survival of CRC patients (45). In addition, the activation of epithelial STAT3 was reported to downregulate the expression of chemokines important for the recruitment of Treg cells in the intestine (24). Therfore, it is tempting to speculate that during *H. pylori* infection, activation of STAT3 in intestinal and colonic epithelial cells contributes to loss of Treg recruitment, thereby supporting malignant transformation of the intestinal tissue. This central role for STAT3 during carcinogenesis in the intestine is supported by the fact that depletion of STAT3 in *Apc*^+/min^ mice led to a reduction in the incidence of early adenomas (46). Importantly, we observed a reduction and normalization of STAT3 levels after eradication of *H. pylori*, which resulted in a normalization of tumor load. Notably, for eradication, a treatment regimen also applied in humans was used in order to be able to translate the results to humans.

The function of STAT3 as oncogene or tumor suppressor seems to be determined by the milieu eliciting its activation as well as the local gut microbiota (47), which is increasingly recognized as an important regulator of colonic cancer development (48). Interestingly, microbial induction of IL-17A production has been shown to endorse colon cancer initiation and progression in *Apc*^+/min^ mice, which was mediated via STAT3 signaling (49). We thus hypothesized that alterations in microbiota compositions in the intestine and colon induced by *H. pylori* may also contribute to carcinogenesis (29, 50). Indeed, when housing mice under germ-free conditions, activation of STAT3 and tumor development were lower upon *H. pylori* infection, but not completely normalized, indicating that microbiota alterations are involved in the phenotype but not exclusively responsible.

Such disturbances in gut microbiota communities have been shown to contribute to CRC development and progression (28). *H. pylori* is known to affect not only local gastric microbiota, but also distant microbial populations in intestine and colon (29, 51). It has been shown that inflammation-driven dysregulation of microbiota can promote colorectal tumor formation and progression (52) and that in response to bacterial stimuli or pathogen associated molecular receptors, pro-inflammatory pathways such as c-Jun/JNK and STAT3 signaling pathways are activated and accelerate intestinal tumor growth in *Apc*^+/min^ mice (47). This was supported by our findings from transferring stool of *H. pylori*-infected SPF mice into germ-free *Apc*^+/1638N^ mice, where we observed an accelerated tumor development in comparison to stool transferred from non-infected mice. Together, these data indicate that *H. pylori* induced microbial signatures are involved in and indispensable to promote intestinal tumor growth.

Our data revealed a distinct mucus-degrading microbiota signature associated with *H. pylori* infection in mice, namely enrichment with *Akkermansia spp.* and *Ruminococcus spp.*, while in human samples from *H. pylori* infected patients, bacterial taxa associated with CRC, Prevotellaceae and Peptostreptococcales, were found (28). Although some studies established an inverse correlation between the presence of *Akkermansia* and gastrointestinal diseases (53), *Akkermansia* has been reported to be increased in CRC patients most likely due to the overexpression of certain mucins in the tumors (54). Notably, we also observed a general loss of goblet cells, which are important to produce mucins and antimicrobial peptides. This loss of goblet cells was also observed in clinical samples from *H. pylori* infected patients undergoing colonoscopy. Thus, *H. pylori* infection disrupts, by two distinct mechanisms, intestinal mucus integrity essential to maintain a healthy barrier to impair bacterial penetration. In the absence of a sufficient regulatory T cell response – as observed here -, which normally keeps inflammatory signals at bay, the carefully balanced homeostasis maintained in the gut by the interplay of a “healthy” microbiome and an intact mucosa then fails to balance the proinflammatory signature elicited by *H. pylori* infection, enabling carcinogenesis. Eradication of *H. pylori* restored intestinal homeostasis with reappearance of goblet cells and normalized the intestinal immune signature, which then completely abrogated the tumor-promoting effect.

Importantly, when analyzing colonic biopsies from *H. pylori* infected patients, we could observe the very same alterations as seen in mice, with activation of pro-carcinogenic signaling pathways and a significant reduction in Treg cells, and an increase of CD3^+^ cells. The attenuated phenotype in eradicated patients highlight the clinical relevance of our findings and indicate, that *H. pylori* infection is more than a mere risk factor for colon carcinogenesis, but actively promotes a procarcinogenic niche in the colon, that may be prevented by eradication of *H. pylori*, which therefore could decrease the risk of CRC development in infected individuals. However, studies showing a correlation between *H. pylori* infection and CRC did not address the effect of antibiotic therapy. The inclusion of such cohorts in future studies is important to determine the impact of *H. pylori* eradication in CRC development.

In summary, our study not only provides solid experimental evidence that *H. pylori* infection accelerates tumor development, but also offers insight into the underlying mechanisms and suggests *H. pylori* screening and eradication as a potential measure for CRC prevention strategies.

## Data Availability

All data produced in the present study have been deposited under BioProject ID PRJNA808836 or are available upon reasonable request to the authors.

http://www.ncbi.nlm.nih.gov/bioproject/808836

## ACKNOWLEDGEMENTS

We thank members of the laboratory ‘Chronic inflammation and carcinogenesis’ for experimental help as well as critical discussion, with special thanks to Maximilian Koch, Karin Taxauer, Martin Skerhut and Teresa Burrell for experimental support. We thank Julia Horstmann and the team of the ColoBac study at the Klinik und Poliklinik für Innere Medizin II, Klinikum rechts der Isar, for the supply of human biopsies. We thank Core Facility Mikrobiom of the ZIEL Institute for Food & Health, Technical University of Munich, for 16S rRNA Sequencing services, as well as Dharmesh Singh and Nyssa Cullin. We thank Core Facility Gnotobiology of the ZIEL Institute for Food & Health, Technical University of Munich, for germ-free mice.

## AUTHOR CONTRIBUTIONS

A.R., R.M.L and M.G. conceived the study. A.R., A.D. and R.M.L. designed and analyzed experiments. S.J. contributed and provided code and support to single cell RNA sequencing and chip cytometry. V.E. and A.W. contributed to experiments. A.D., S.J, and R.M.L contributed to data interpretation. M.V. and M.Q. provided human biopsies. K.P.J. provided mouse models and critically revised the article. D.H., D.H.B. and L.D. critically revised the article. A.R. and R.M.L. wrote the article. A.R., R.M.L. and M.G. revised the article. M.G. acquired the funding. All authors read and reviewed the article.

## DATA AVAILABILITY

Raw single cell RNA sequencing and 16S rRNA sequencing data are available under the BioProject ID PRJNA808836.

## MATERIALS AND METHODS

### STUDY DESIGN

This study was conceptualized to investigate the underlying mechanisms of *H. pylori* induced colorectal carcinogenesis.

Tumor mouse models *Apc*^+/min^, initially obtained from Jackson Laboratories, and *Apc*^+/1638N^ mice, provided by Prof. Klaus-Peter Janssen (Klinikum rechts der Isar, München) (41), were bred under specific pathogen-free conditions at our animal facility at the Technical University of Munich. Both female and male mice were used and co-housed with littermate controls.

*Apc*^+/1638N^ mice and wild type littermates were re-derived germ-free from conventional mice by Prof. Bleich and Dr. Basic (Hannover medical school, Hannover).

Female C57BL/6 mice were purchased from Envigo RMS GmbH at an age of 6 weeks and acclimatized to our animal facility for 1-2 weeks prior infection. Mice were fed with a standard diet and water ad libitum and maintained under a 12-hour light-dark cycle. DNA extracted from mouse tails was used for genotyping. All animal experiments were conducted in compliance with European guidelines for the care and use of laboratory animals and were approved by the Bavarian Government (Regierung von Oberbayern, Az.55.2-1-54-2532-161-2017).

87 fresh colonoscopy biopsies were collected within the framework of the COLOBAC study of the CRC1371 (Dept. of Surgery and II. Medical Dept. Klinikum rechts der Isar, Technical University of Munich, Germany). 67 FFPE colon biopsies were obtained from the Klinikum Bayreuth. Both studies were approved by the respective ethics committees (Klinikum rechts der Isar #322/18, Klinikum Bayreuth #241_20Bc).

*H. pylori* status of colonoscopy biopsies was determined in serum samples using the recomwell Helicobacter IgG kit (Mikrogen) according to manufacturer’s instructions. *H. pylori* status of FFPE biopsies was determined histologically in corresponding gastric biopsies by Prof. Michael Vieth.

### RANDOMISATION AND BLINDING

Animals were randomly distributed to control/infection groups. The infection status of mice was known to the investigating researcher. *H. pylori* status of patients was not known to the researcher during sample analysis.

### H. PYLORI INFECTION

*H. pylori* pre-mouse Sidney Strain 1 (PMSS1) was cultured on Wilkins-Chalgren (WC) Dent (containing vancomycin, trimethoprim, cefsoludin and amphotericin) agar plates in a microaerophilic atmosphere (5% O2, 10% CO2). 6–8-week old mice were orally gavaged twice within 72 hours with 2 × 10^8^ *H. pylori* PMSS1 in 200 µl brain-heart-infusion (BHI) medium containing 20% fetal calve serum (FCS). Infection status was determined by plating homogenized stomach tissue on WC Dent plates supplemented with 200 g/ml bacitracin, 10 g/ml nalidixic acid and 3 g/ml polymycin B, and counting colony-forming units (CFU).

### *H. PYLORI* ERADICATION

After 4 weeks of infection, *H. pylori* eradication was performed with an antibiotic cocktail containing clarithromycin (Eberth) (7.15 mg/kg/day), metronidazole (Carl Roth) (14.2 mg/kg/day) and the proton-pump inhibitor omeprazole (Carl Roth) (400 µmol/kg/day) by oral gavage twice daily for 7 consecutive days. Omeprazole was dissolved in 200µl 2.5% Hydroxy-propyl-methyl-cellulose (Sigma-Aldrich) with pH adjusted to 9. Antibiotics, dissolved in 200µl PBS, were administered 45 minutes after omeprazole (30).

### STOOL TRANSFER

SPF *Apc*^+/1638N^ mice and wild type littermates were infected at an age of 6-8 weeks and after 24 weeks of infection, stool pellets were collected from these “donor” mice, dissolved in 0.1ml PBS/g stool and administered via oral gavage to germ-free *Apc*^+/1638N^ “recipient” mice.

### HISTOLOGY AND IMMUNOHISTOCHEMISTRY

Small intestine and colon were longitudinally opened and flushed with phosphate-buffered saline (PBS). Solid neoplastic lesions were assessed and measured macroscopically by two independent examiners. Dissected tissue was fixed in 4% formaldehyde, embedded in paraffin and 4µm thick sections were used for staining. For immunohistochemical stainings, antigen retrieval was achieved with 10 mM sodium citrate (pH 6) or 1 mM EDTA (pH 8), and primary antibodies were applied overnight at 4°C (Table 1). Horseradish peroxidase (HRP) coupled secondary antibodies (Promega) and diaminobenzidine (DAB) (CellSignaling) were used to detect signal. Periodic acid Schiff (PAS) (Carl Roth) staining was performed to assess the quantity of mucus producing goblet cells. Stomach, intestinal and colonic sections were blindly quantified by two independent researchers by measuring the area of a functional unit (stomach gland, intestinal crypt/villus unit or colonic crypt) and counting positive cells per mm^2^, using Aperio ImageScope (Leica BioSystems).

**Table 1.** Antibodies used for immunohistochemical evaluation.

| Target | Clone | Origin/Target | Antigen retrieval | Dilution | Company |
| --- | --- | --- | --- | --- | --- |
| CD3 | SP7 | Rabbit mAB | sodium citrate | 1:150 | Thermo Fisher |
| Ki67 | D3B5 | Rabbit mAB | sodium citrate | 1:400 | Cell Signaling |
| pSTAT3 | D3A7 | Rabbit mAB | EDTA | 1:200 | Cell Signaling |

### CHIPCYTOMETRY AND AUTOMATIC IMAGE QUANTIFICATION

Murine intestinal and colonic cross-sections were preserved with O.C.T. compound (Tissue Tek) in cryomolds (Tissue Tek) and kept frozen at −80°C. For ChipCytometry, 7µm thick sections were cut on a Cryostat (Leica), fixed in Fixation Buffer (ZELLKRAFTWERK) for 45 minutes and subsequently transferred to CellSafe Chips (ZELLKRAFTWERK). ChipCytometry on human FFPE biopsies was performed according to the procedure described in Jarosch, Köhlen et al (55). Briefly, tissue sections were rehydrated on coverslips and antigen retrieval was performed using TRIS-EDTA buffer (pH 8.5) and then transferred to CellSafe Chips (ZELLKRAFTWERK). Alternating cycles of staining, immunofluorescence detection and photobleaching were performed for various markers (Table 2). Automated image processing was performed as described in Jarosch, Köhlen et al., which includes segmentation of cells, removing of outliers and spatial spill over correction (55). The resulting cell – marker matrix was analyzed using FlowJo software (V10.8.0), which enabled absolute quantification of cells.

**Table 2.** Antibodies used for ChipCytometry.

| Epitope | Fluorochrome | Clone | Dilution | Company | Catalog # |
| --- | --- | --- | --- | --- | --- |
| anti-mouse CD3 | PerCP/Cy5.5 | 17A2 | 1:200 | BioLegend | 100218 |
| anti-mouse CD4 | eFluor450 | RM4-5 | 1:80 | BioLegend | 100531 |
| anti-mouse CD45 | FITC | 30-F11 | 1:100 | BioLegend | 103108 |
| anti-mouse Foxp3 | PE | FJK-16s | 1:80 | eBioscience | 14-5773-82 |
| Pan-cytokeratin | AlexaFluor 488 | C-11 | 1:100 | BioLegend | 628602 |
| Hoechst | BUV395 | - | 1:50.000 | ThermoScientific | H3570 |
| α-SMA | eFluor570 | 1A4 | 1:750 | eBioscience | 41-9760-80 |
| anti-human CD3 | unconjugated | SP7 | 1:150 | ThermoScientific | RM-9107-S1 |
| anti-human CD4 | AlexaFluor 488 | polyclonal | 1:50 | R&D Systems | FAB8165G |
| anti-human CD45 | PerCP/Cy5.5 | HI30 | 1:80 | BioLegend | 304028 |
| anti-human Foxp3 | PE | 236A/E7 | 1:30 | eBioscience | 563791 |
| 2<sup>nd</sup> anti-rabbit | PE | Polyclonal | 1:300 | BioLegend | 406421 |

### LAMINA PROPRIA AND INTRAEPITHELIAL LYMPHOCYTE ISOLATION AND FLOW CYTOMETRY

Harvested intestinal tissue was cut open longitudinally after removing Peyer’s Patches and adjacent tissue. Subsequently, tissue was treated with 30mM EDTA, filtered supernatants were collected as intraepithelial lymphocytes and remaining tissue was digested with 0.5mg/mL collagenase from Clostridium histolyticum Type IV (Sigma Aldrich) and 10μg/mL DNase I (Applichem). Filtered and centrifuged lamina propria cell suspensions were density separated using a Percoll gradient (Thermo Fisher).

Fresh human biopsies were collected in Hank’s Balanced Salt Solution w/o Mg^2+^/Ca^2+^ (HBSS) and digested with 0.1% collagenase from Clostridium histolyticum Type IV (Sigma Aldrich) for 30 min. at 37°C. Digestion was stopped by adding 20 mL HBSS and centrifugation twice. Isolated lymphocytes were frozen in Dimethyl sulfoxide (Applichem) + 20 % FCS at −80° C.

Single cell suspensions were blocked with anti-mouse CD16/CD32 or anti human TruStain FcX and live/dead staining performed with Zombie Aqua (BioLegend) in PBS. Surface antibodies (Table 3) were diluted according to titration experiments and cells stained for 30 min at 4° C. For transcription factors, Foxp3 Transcription Factor Staining Buffer Set (eBioscience) was used according to manufacturer’s instructions. For stimulation with whole *H. pylori* lysate, cells were stimulated for 12 hours with 20µg/mL PMSS1 lysate at 37° C and protein transport inhibitor Golgi Plug (BD Biosciences) added 1:1000 after 7 hours for a total of 5 hours. Stimulated cells were stained with intracellular cytokine staining kit according manufacturer’s instructions (BD Biosciences). Stained single cell suspensions were acquired on a CytoFlex S (Beckman Coulter) and analyzed using FlowJo software (V10.8.0).

**Table 3.** Antibodies used for flow cytometry.

| Epitope | Fluorochrome | Clone | Dilution | Catalog # | Company |
| --- | --- | --- | --- | --- | --- |
| anti-mouse CD16/CD32 | - | 93 | 1:500 | 14-0161-86 | eBioscience |
| anti-mouse CD45 | AlexaFluor 700 | 30-F11 | 1:400 | 103128 | BioLegend |
| anti-mouse CD3ε | FITC | 500A2 | 1:200 | 152304 | BioLegend |
| anit-mouse CD4 | BV605 | RM4-5 | 1:250 | 100548 | BioLegend |
| anti-mouse CD4 | eFluor450 | RM4-5 | 1:250 | 48-0042-82 | eBioscience |
| anti-mouse CD8a | APC-H7 | 53.6-7 | 1:250 | 560182 | BD Biosciences |
| anti-mouse FoxP3 | eFluor450 | FJKL-16s | 1:200 | 45-5773-82 | eBioscience |
| anti-mouse IL-17A | APC | eBio17B7 | 1:150 | 17-7177-81 | eBioscience |
| anti-human CD45 | AlexaFluor 700 | 2D1 | 1:50 | 368514 | BioLegend |
| anti-human CD3 | FITC | OKT3 | 1:300 | 317306 | BioLegend |
| anti-human CD4 | PB450 | SK3 | 1:30 | 344620 | BioLegend |
| anti-human CD8 | APC-H7 | SK1 (RUO) | 1:150 | 560179 | BD Biosciences |
| anti-human FoxP3 | PerCP/Cy5.5 | PCH101 |  | 35-4776-42 | eBioscience |
| Human TruStain FcX | - | - | 1:500 | 422302 | BioLegend |

### SINGLE CELL RNA SEQUENCING

Intestinal and colonic tissue was harvested and cells were isolated as described in section “Lymphocyte isolation and Flow cytometry”. Single cell suspensions were stained with anti-mouse CD45 PB450 (Clone: 30-F11, BioLegend, #103126), anti-mouse EPCAM APC (Clone: G8.8, BioLegend, #118214) and Propidium Iodide (PI). CD45+ PI- and EPCAM+ PI- cells were sorted.

For cell hashing TotalSeq-B anti-mouse Hashtags 1, 2 and 5 to 8 (M1/42; 30-F11, Biolegend, 155831, 155833, 155839, 155841, 155843, 155845) were used at a dilution of 1:50.

Single cell RNA Sequencing was performed with 10X Genomics, according to manufacturer’s instructions (Chromium™ Single Cell 3’ GEM v3 kit). Sorted cells were centrifuged and resuspended in mastermix and 37.8 µl of water, before 70 µl of the cell suspension was transferred to the chip. QC was performed with a high sensitivity DNA Kit (Agilent) on a Bioanalyzer 2100, and libraries were quantified with the Qubit dsDNA HS assay kit (life technologies).

Libraries were pooled according to their minimal required read counts (20.000 reads/cell for gene expression libraries). Illumina paired end sequencing was performed with 150 cycles on a NovaSeq 6000.

Annotation was performed using cellranger (V5.0.0, 10X genomics) against the murine reference genome GRCm38 (mm10-2020-A). All subsequent analysis was performed using SCANPY V1.6 (56). Preprocessing was performed following the guidelines of best practice in single-cell RNA-seq analysis (57) and involved less than 15% mitochondrial genes, regressing out cell cycle, mitochondrial genes and total counts. The data was normalized per cell count and logarithmized. Genes used for gene scores are listed in Table S1 and the scores were computed with the SCANPY build in function “sc.tl.score_genes”.

RNA velocities were calculated using velocyto (18) and analyzed with scVelo (V 0.2.3) (58).

### 16S RRNA SEQUENCING

Bacterial DNA extraction and 16S rRNA sequencing was either performed as described previously (59, 60) by the Core Facility Microbiome of the ZIEL Institute for Food & Health (Technical University of Munich) or as follows: small intestine and colon tissue were homogenized with a Precellys® 24 homogeniser (Avantor). Phenol chloroform DNA isolation and ethanol precipitation were performed following modified protocols of P.J. Turnbaugh et al., 2009 and E. G. Zoetendal et al. 2006 (61, 62). Subsequently, the V3/V4 region of the *16S rRNA* gene was amplified and double indexed using barcoding primers modeled after Kozich *et al*. PCR fragments were purified using magnetic AMPure XP beads (Beckman Coulter, USA) according to manufacturer’s instructions (63). The final pooled library was sequenced on an Illumina MiSeq with Reagent Kit v3 (Ilumina) for 600 cycles of paired-end reads.

Raw sequences were analyzed using the Qiime2 platform (v2021.4) (64). In detail, denoising, removing of chimeras and generation of Amplicon Sequence Variants (ASVs) was performed with dada2. Subsequently, a phylogenetic tree was generated and diversity measures were calculated. Chao1 index was used to determine community alpha diversity. Taxonomic classification was performed with a qiime2 feature classifier trained on the SILVA132 99% OTUs, specifically targeting the V3 region. Linear discriminant analysis effect size (LEfSe) determining differentially abundant features was performed on the online interface at http://huttenhower.sph.harvard.edu/lefse/, developed by Segata et al. (65).

### QUANTITATIVE PCR

Stomach, small intestine and colon tissue were homogenized with Precellys® 24 homogeniser (Avantor) and RNA isolation was performed with a Maxwell 48 RSC simply RNA Tissue Kit on a Maxwell RSC Instrument (Promega). cDNA was synthesized with Moloney Murine Leukemia Virus Reverse Transcriptase RNase H-Point Mutant (Promega). Gene expression was assessed with GoTaq qPCR Mastermix (Promega) on a CFX384 system (Bio-Rad). The quantitative PCR consisted of 40 cycles of amplification with 15 sec denaturation at 95 ^◦^C, 1 min annealing and amplification at 60 ^◦^C. According to the ΔΔCT method, CT values were normalized to *Gapdh* and to uninfected controls, in order to determine fold changes in gene expression. The sequences of primers used are summarized in Table 3.

**Table 3.**
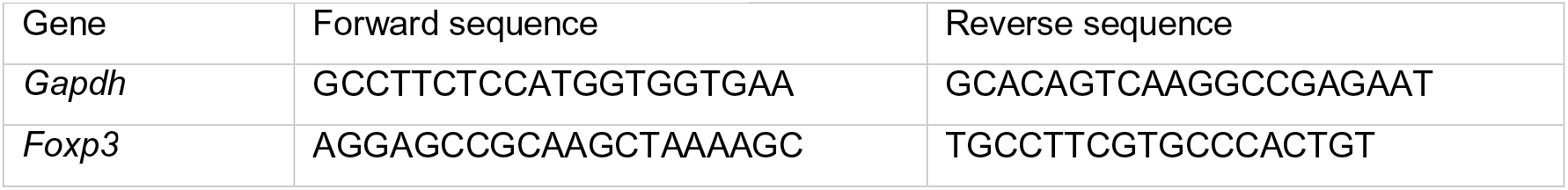
Primer sequences used for qPCR.

### STATISTICAL ANALYSIS

Statistical analysis was conducted on biological replicates as stated in the Fig. legends. Depending on Gaussian distribution, statistical significance between two groups was determined with unpaired student’s t-test or Mann-Whitney-U test and for analysis among more than two groups, one-way ANOVA with Tukey’s multiple-comparisons test or Kruskal-Wallis-test with Dunn’s multiple-comparisons test. P values below 0.05 were considered significant. Exact p values are stated when relevant. Statistical analysis was carried out using Prism 8 (GraphPad Software).

---

**Fig. S1:**
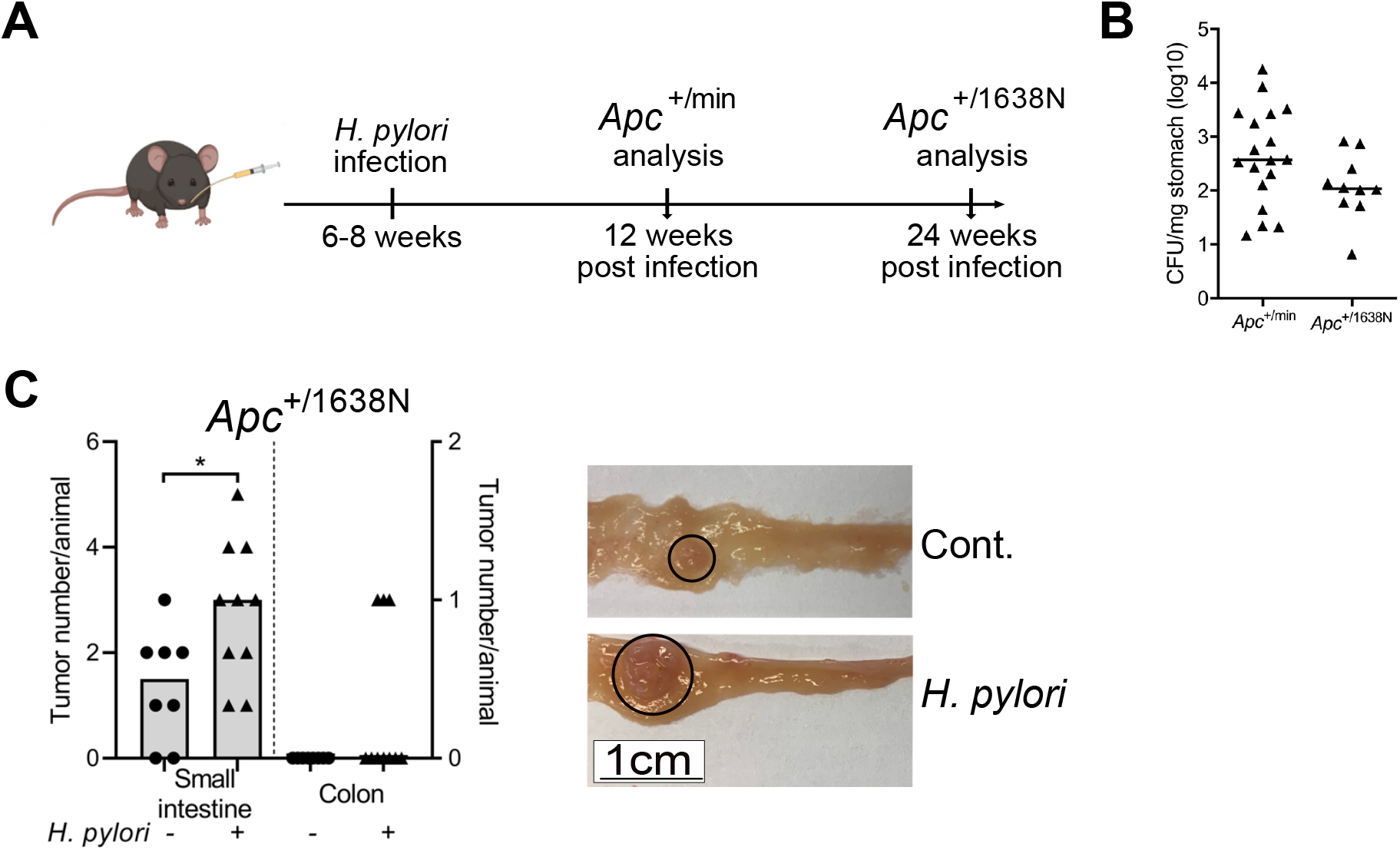
(A) Experimental setup for infection of *Apc*^+/min^ and *Apc*^+/1638N^ mice. (B) Colony forming units (CFU) per milligram (mg) stomach tissue of *H. pylori* infected *Apc*^+/min^ mice after 12 weeks of infection and *Apc*^+/1638N^ mice after 24 weeks of infection. (C) Tumor counts of *H. pylori* infected (n=10) and non-infected (n=8) *Apc*^+/1638N^ mice after 24 weeks of infection in small intestine and colon. Representative pictures showing tumors (circle) of non-infected (Cont.) and infected (*H. pylori*) mice. Each symbol represents one animal, from 2-3 independent, pooled experiments. Bars denote median. Statistical significance was determined with Mann-Whitney-U test or unpaired t-test, *p < 0.05.

**Fig. S2:**
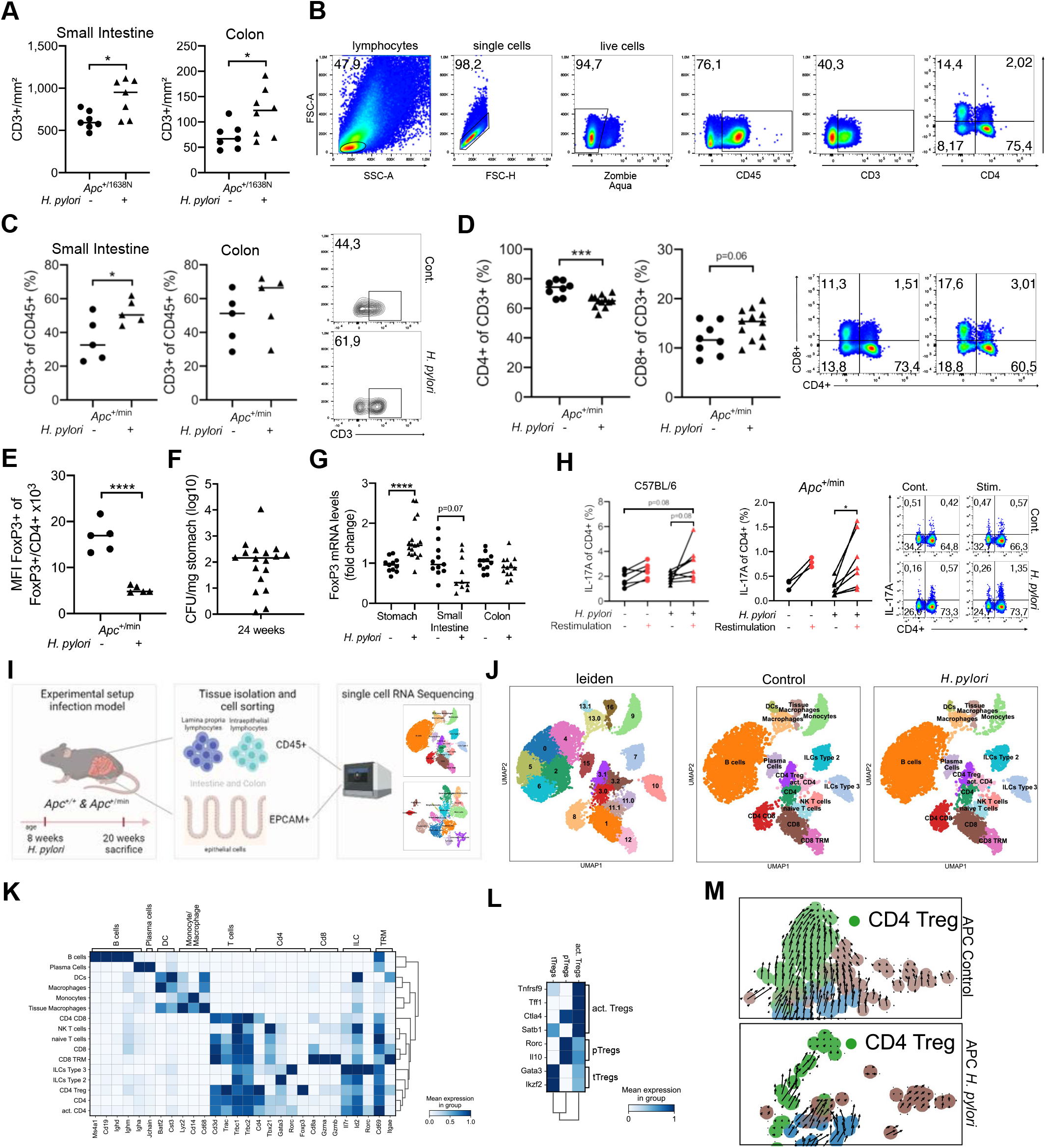
(A) Quantification of CD3+ cells per mm^2^ small intestine and colon tissue is shown of *H. pylori* infected and non-infected *Apc*^+/1638N^ mice. Pooled data of 3 independent experiments. (B) Gating strategy of intestinal lymphocytes. Preliminary lymphocyte gate was applied based on known size and granularity on forward and sideward scatter, followed by exclusion of doublets and dead cells. Afterwards, CD45+ lymphocytes and CD3+ T-cells were selected and subsequent gating was performed on CD4+ or CD8+ T-cells. (C) Frequencies of intraepithelial T-cells detected by flow cytometry in small small intestine and colon of *H. pylori* infected and non-infected *Apc*^+/min^ mice, gated on live, single cells and CD45+. Representative contour plots are included. Data of one representative experiment of 3 independent experiments is shown. (D) Frequencies of T-cell subtypes detected by flow cytometry of *H. pylori* infected and non-infected *Apc*^+/min^ mice, gated on live, single cells, CD45+ and CD3+. Representative pseudocolor plots are included. Pooled data of two independent experiments are shown. (E) Mean fluorescence intensity (MFI) of FoxP3+ CD4+ T-cells detected by flow cytometry of *H. pylori* infected and non-infected *Apc*^+/min^ mice, gated on live, single cells, CD45+, CD3+ and CD4+. Data of one representative experiment of 3 independent experiments is shown. (F) Colony forming units (CFU) in homogenized stomach tissue of *H. pylori* infected C57BL/6 mice after 24 weeks of infection. Pooled data of 2 independent experiments are shown. (G) Fold change in Foxp3 mRNA expression in stomach, small intestine and colon tissue homogenates, normalized to Gapdh and non-infected’control mice (2-ΔΔCT value). Pooled data of 2 independent experiments. (H) Flow cytometric analysis of intestinal lamina propria lymphocytes isolated from *H. pylori* infected and non-infected *Apc*^+/min^ mice. Paired frequency of IL-17A positive cells of CD4+ T-cells (non-stimulated vs. stimulated with whole *H. pylori* lysate) gated on live, single cells, CD45+ and CD3+, and representative pseudocolor plots of IL-17 releasing CD4+ T-cells are shown. Data of one representative experiment of 2 independent experiments is shown. (I) Experimental setup of single-cell RNA sequencing of CD45+ and EPCAM+ cells isolated from small intestine and colon of *H. pylori* infected and non-infected *Apc*^+/min^ mice and *Apc*^+/+^ mice. 2 mice per group were pooled into one sample. (J) Unsupervised clustering of immune cells plotted as uniform manifold approximation and projection (UMAP), n=2 mice per group, n= 11407 cells. Leiden clustering and annotated clusters for non-infected and infected mice are shown. (K) Gene matrix of marker genes used for annotation of CD45+ clusters. (L) Gene matrix of marker genes used for annotation of Treg subclusters. (M) RNA velocity analysis plotted as UMAP for single CD3+ T cells from non-infected (left) and *H. pylori* infected *Apc*^+/min^ mice (right). The directional flow of the velocity arrows between cell clusters shows the projection from the observed state to the predicted future state. Zoom into CD4 Treg RNA velocity analysis of *H. pylori* infected and non infected *Apc*^+/min^ mice Each symbol represents one animal/single cell, pooled from at least 2 independent experiments (n=6-10 mice/group/experiment) or 2 mice/group for single-cell data. Bars denote median. Unless otherwise specified, statistical significance was determined with student’s t-test in case of normal distribution, otherwise by Mann-Whitney U test, *p < 0.05, **p < 0.01, ***p < 0.001, ****p < 0.0001.

**Fig. S3:**
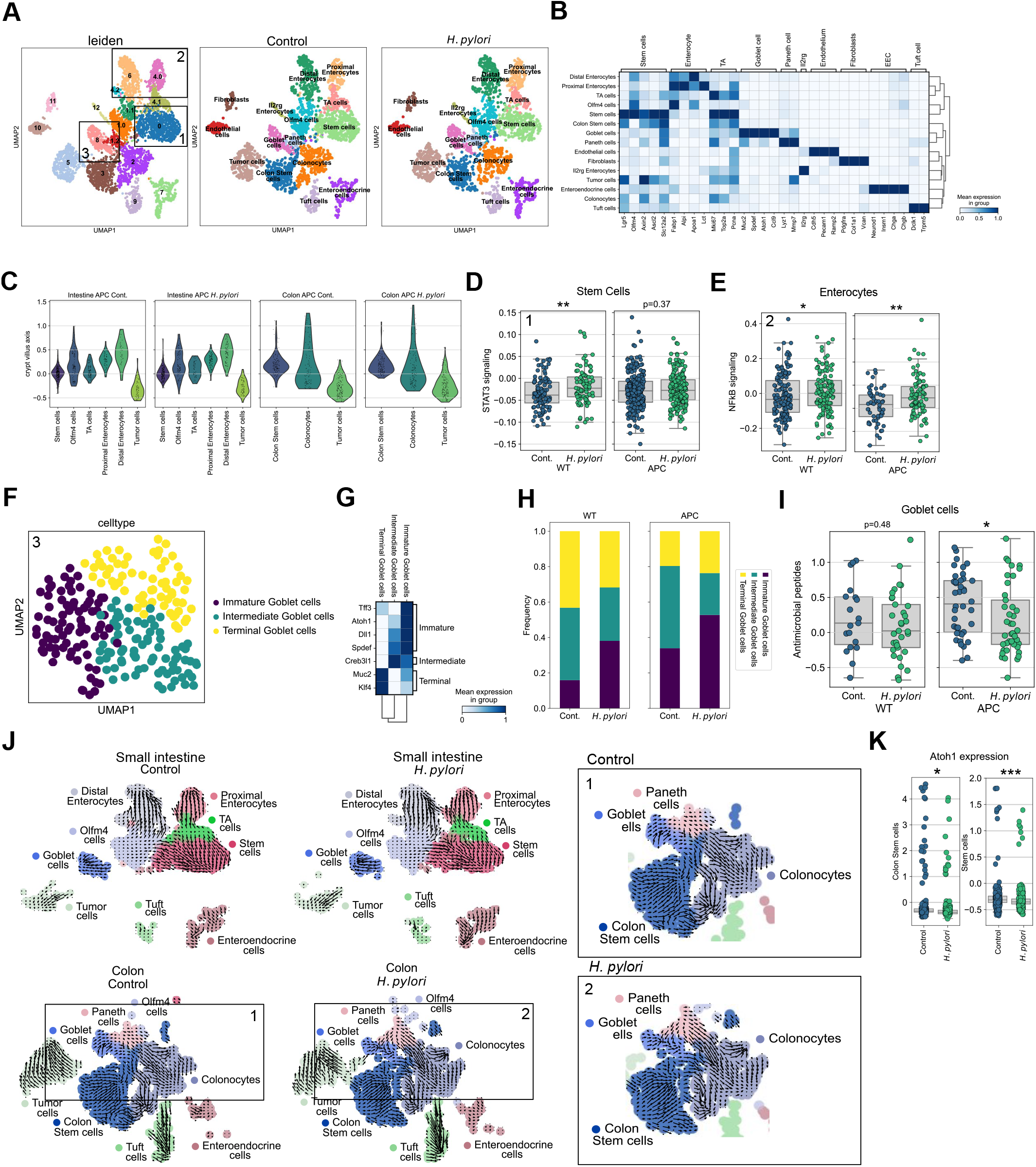
(A) Unsupervised clustering of epithelial cells plotted as uniform manifold approximation and projection (UMAP), n=2 mice per group, n= 4249 cells. Leiden clustering and annotated clusters for non-infected and infected mice are shown. Clusters for further analysis are highlighted (1= Stem Cells, 2=Enterocytes, 3= goblet cells). (B) Gene matrix of marker genes used for annotation of EPCAM+ clusters. (C) Pseudo-spatial distribution of stem-cell, enterocyte and tumor cell clusters along the crypt villus axis in small intestine and colon of *Apc*^+/+^ (WT) and *Apc*^+/min^ (APC) mice. (D) Gene set score of STAT3 signaling genes, comparing intestinal stem cells from *H. pylori* infected and non-infected *Apc*^+/+^ (WT) and *Apc*^+/min^ (APC) mice. (E) Gene set score of NFkB signaling genes, comparing intestinal enterocytes from *H. pylori* infected and non-infected *Apc*^+/+^ (WT) and *Apc*^+/min^ (APC) mice. (F) Unsupervised clustering and reannotation of goblet cluster plotted as UMAP, n=218 cells. (G) Gene matrix of marker genes used for annotation of goblet cell subclusters. (H) Relative frequency of goblet subtypes in *Apc*^+/+^ (WT) and *Apc*^+/min^ (APC) mice, grouped by infection. (I) Gene set score of antimicrobial peptide genes, comparing immature goblet cells of small intestine from *H. pylori* infected and non-infected *Apc*^+/+^ (WT) and *Apc*^+/min^ (APC) mice. (J) RNA velocity analysis plotted as UMAP for single EPCAM+ cells from non-infected and *H. pylori* infected mice in small intestine and colon. The directional flow of the velocity arrows between cell clusters shows the projection from the observed state to the predicted future state. Squares highlight and zoom in to goblet, paneth cell and coloncytes clusters from non-infected (=1) and infected (=2) colonic epithelial cells. (K) Expression of goblet cell differentiation marker Atoh1 in Colon Stem cells and small intestinal Stem cells, comparing *H. pylori* infected and non-infected mice. Each symbol represents one single-cell, pooled from 2 mice/group. Bars denote median. Statistical significance was determined with Kruskal-Wallis test, *p < 0.05, **p < 0.01, ***p < 0.001.

**Fig. S4:**
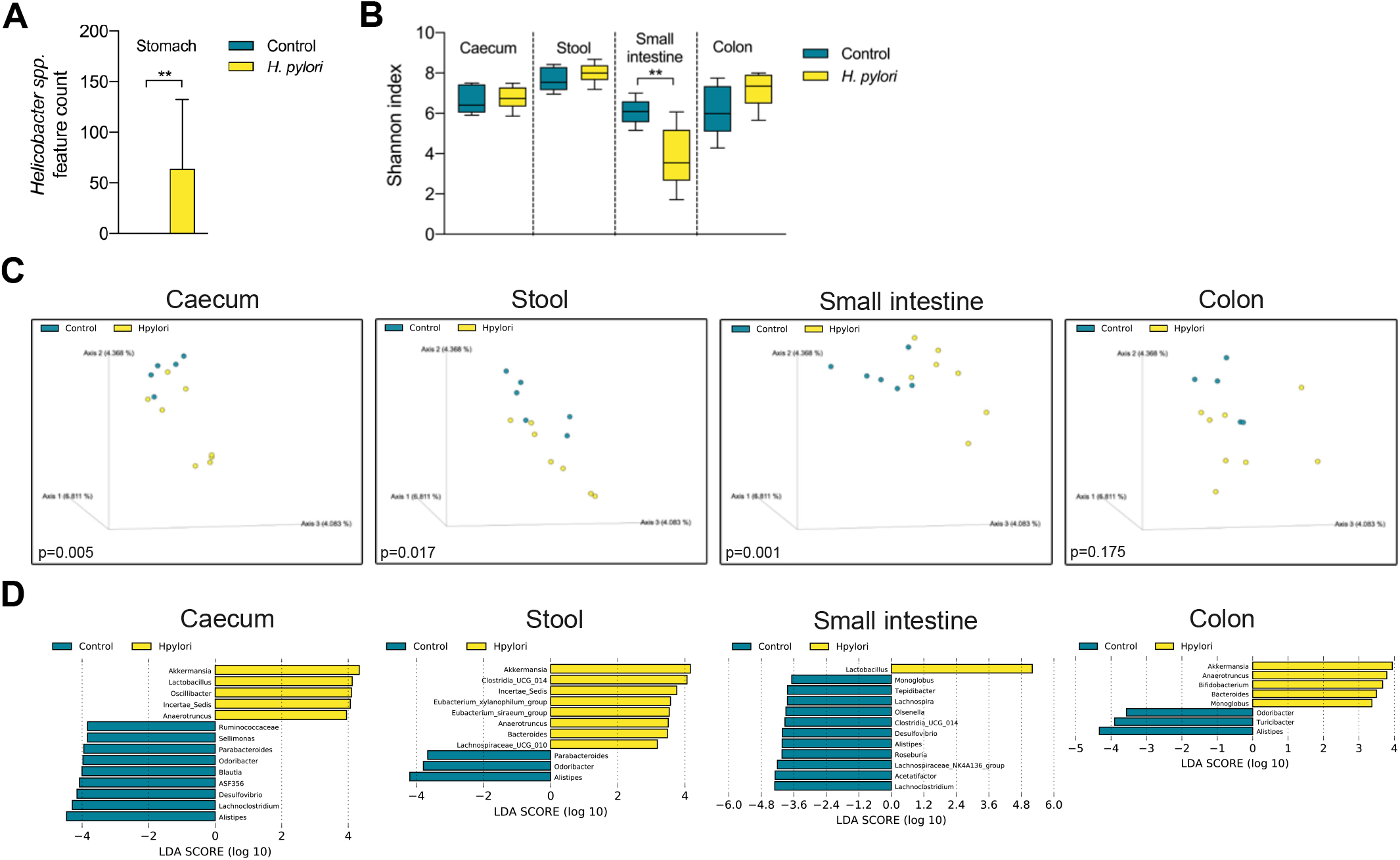
(A) Feature counts (ASVs) of *Helicobacter spp.* in stomach of *H. pylori* infected and non-infected C57BL/6 mice (n=6-8mice/group). Data of one representative experiment of 2 independent experiments. Shown as bars with mean and standard deviation (SD). Statistical significance was determined with Mann-Whitney U test, **p < 0.01. (B) Shannon index depicting alpha-diversity in ceacum, stool, small intestine and colon of *H. pylori* infected and non-infected C57BL/6 mice (n=6-8mice/group). Data of one representative experiment of 2 independent experiments. Shown as box and whiskers with (SD). Statistical significance was determined with Mann-Whitney U test, **p < 0.01. (C) Bray-Curtis dissimilarity depicting beta-diversity of ceacum, stool, small intestine and colon between *H. pylori* infected and non-infected C57BL/6 mice (n=6-8mice/group). Data of one representative experiment of 2 independent experiments. Statistical significance was determined with PERMANOVA. (D) Linear discriminant effect size analysis (LEfSe) determining differentially abundant features of caecum, stool, small intestine and colon tissue upon *H. pylori* infection of C57BL/6 mice.

**Fig. S5:**
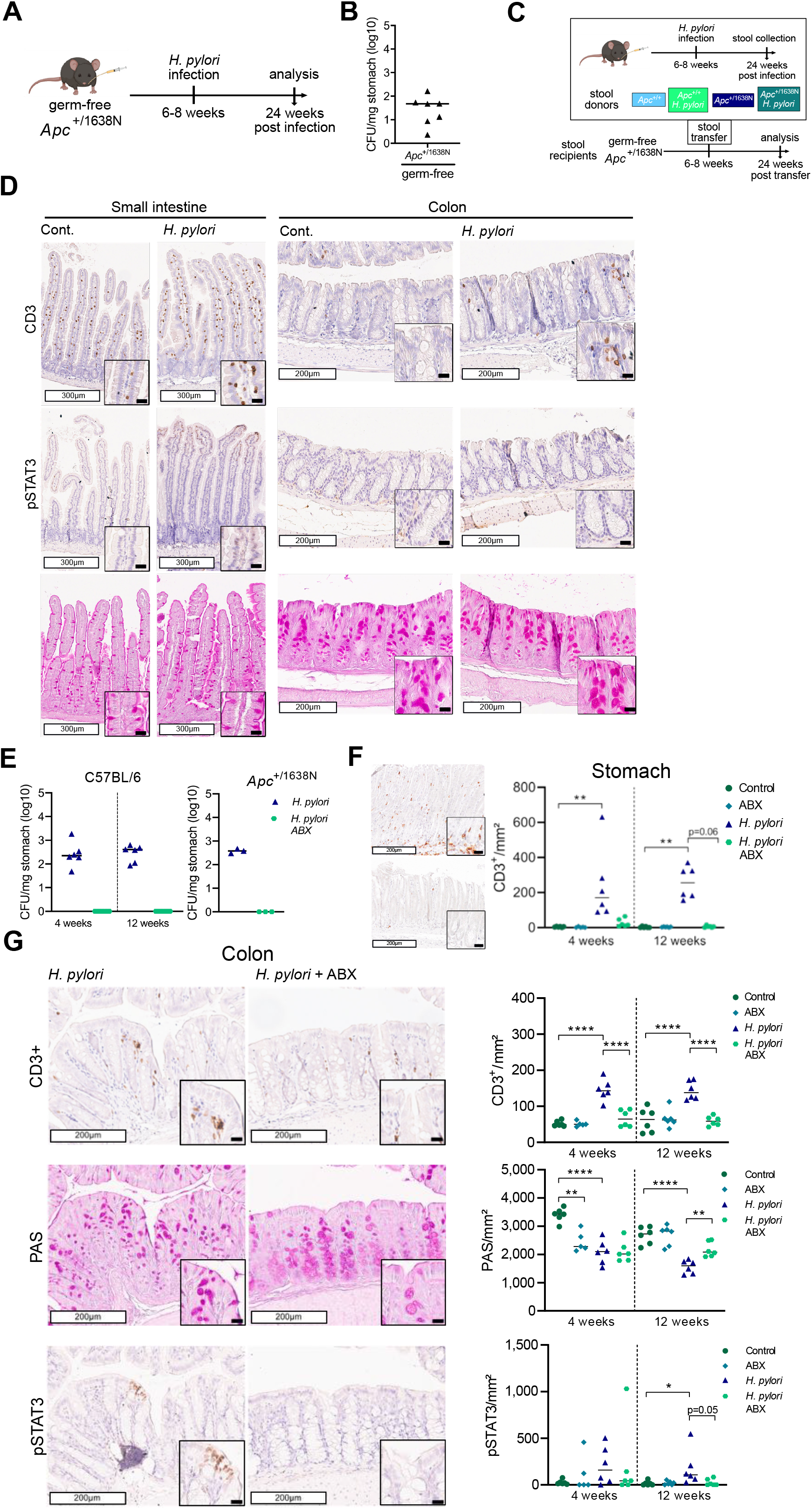
(A) Experimental setup for infection of germ-free *Apc*^+/1638N^ mice. (B) Colony forming units (CFU) per milligram (mg) stomach tissue of H. pylori infected germ-free *Apc*^+/1638N^ mice. Pooled data of two independent experiments (n= 8 mice/group). (C) Experimental setup for stool transfer from specific-pathogen free mice (non-infected and *H. pylori* infected *Apc*^+/+^ or *Apc*^+/1638N^ mice, respectively, (stool donors)) to germ-free *Apc*^+/1638N^ mice (stool recipients). (D) Representative pictures of CD3+, pSTAT3 and PAS stainings from small intestine and colon of non-infected and *H. pylori* germ-free *Apc*^+/1638N^ mice are shown. White scale bars correspond to 300 μm in case of small intestine and 200 μm in case of colon, black scale bars to 20 μm. (E) CFU per mg stomach tissue of *H. pylori* infected and eradicated C57BL/6 and *Apc*^+/1638N^ mice is shown. Data of one experiment (n=3-6 mice/group). (F) Representative pictures of gastric CD3+ staining and quantification of positive intraepithelial cells per mm^2^ of *H. pylori* infected and eradicated C57BL/6 mice are shown. White scale bars correspond to 200 μm, black scale bars to 20 μm. Data of one experiment (n= 5-6 mice/group). (G) Representative pictures of colonic CD3+, pSTAT3 and PAS stainings from H. pylori infected and eradicated C57BL/6 mice are shown. White scale bars correspond to 200 μm, black scale bars to 20 μm. Quantification of positive cells per mm^2^ is shown. Data of one experiment (n= 5-6 mice/group). Each symbol represents one animal. Bars denote median. Statistical significance was determined with ordinary one-way ANOVA with Tukey’s multiple comparisons test in case of normal distribution, otherwise by Kruskal-Wallis-Test with Dunn’s multiple comparisons test, *p < 0.05, **p < 0.01, ***p < 0.001, ****p < 0.0001.

**Fig. S6:**
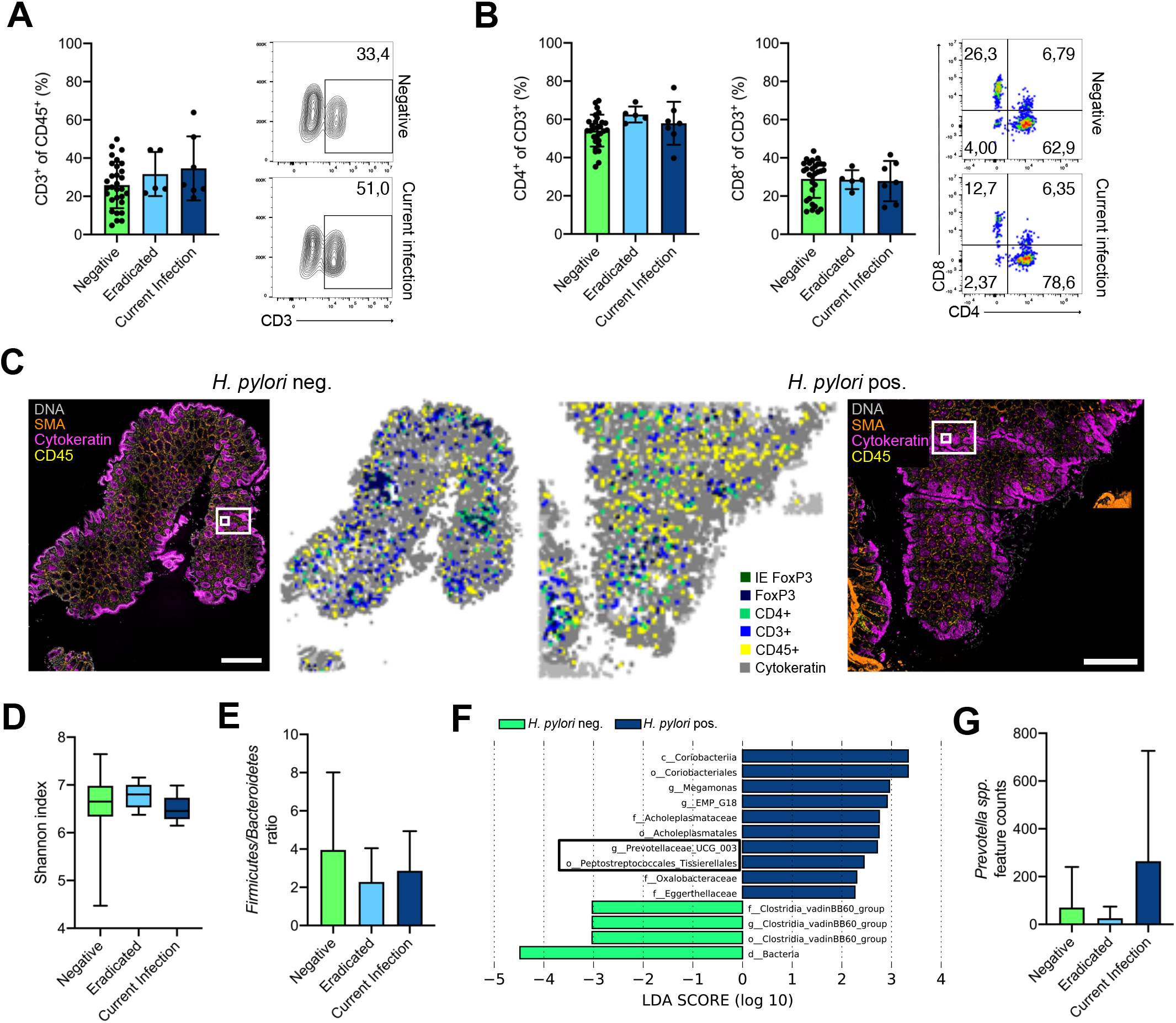
(A) Flow cytometric analysis of colon biopsies from *H. pylori* currently infected, eradicated and non-infected patients were conducted. Frequencies of CD3+ T-cells, gated on live, single cells, CD45+ and CD3+ are shown and representative contour plots of *H. pylori* currently infected and negative individuals are included. (B) Frequencies of T-cell subsets CD4 and CD8, gated on live, single cells, CD45+ and CD3+ are shown and representative pseudocolor plots of *H. pylori* currently infected and negative individuals are included. (C) Overview of colon tissue stained with multiplexed chip cytometry. Automatic image processing of multiplexed chip cytometry on human colon tissue determines CD45+, CD3+, CD4+ and FoxP3+ cells and their location. Large scale bar corresponds to 500μm. (D) Shannon index as indicator of alpha-diversity of stool samples from *H. pylori* currently infected, eradicated and non-infected individuals are shown. Data shown as box and whiskers with SD. (E) Ratio of the phyla Firmicutes to Bacteroidetes, shown as bars with mean and SD. (F) Linear discriminant effect size analysis (LEfSe) determining differentially abundant features of stool samples from *H. pylori* negative and positive individuals. Interesting features are highlighted in black. (G) Feature counts of Prevotella spp. of stool samples *H. pylori* currently infected, eradicated and non-infected individuals, shown as bars with mean and SD. Each symbol represents one patient. Statistical significance was determined with ordinary one-way ANOVA with Tukey’s multiple comparisons test in case of normal distribution, otherwise by Kruskal-Wallis-Test with Dunn’s multiple comparisons test.

